# Unraveling the Lifespan Trajectory of White Matter Hypointensity on T1-weighted MR Images and Clinical Application in Various Brain Disorders

**DOI:** 10.64898/2026.11.12.26367778

**Authors:** Zhizheng Zhuo, Shan Lv, Peng Gao, Ningnannan Zhang, Li Chai, Xiaozhou Du, Yuna Li, Yinshan Wang, Ye He, Ping Wang, Renlong Zhang, James H. Cole, Zixuan Lin, Dennis M. Hedderich, Dan Cheng, Shuiwei Xia, Yunyun Duan, Wei Wang, Yunling Wang, Jiansong Ji, Xi-Nian Zuo, Yaou Liu, Chinese Brain Health Imaging Consortium (CHART)

## Abstract

White matter hypointensity on T1-weighted MRI (T1-WMHypo), a putative marker of white matter pathophysiological changes, has been less systematically investigated than the widely studied T2-weighted fluid-attenuated inversion recovery (T2-FLAIR) hyperintensity lesions. Here, we built the lifespan normative reference and assessed the clinical significance of T1-WMHypo across various brain disorders using a large multicenter dataset. We first established lifespan normative reference for T1-WMHypo volume by 34,148 healthy individuals (aged 4-85 years). Then, in 7,587 patients across 21 neurological and psychiatric disorders, normative reference-derived deviations revealed disease-specific patterns, with marked deviations in neurological diseases. Deviations of T1-WMHypo volume contributed to differential diagnosis, larger deviations correlated with brain atrophy, cognitive impairment, and poorer outcomes in various brain disorders. Elevated burden of T1-WMHypo volume predicted poor medication response in Parkinson’s disease and accelerated disability progression in multiple sclerosis. These associations persisted after adjusting for conventional T2-FLAIR hyperintensity lesions. T1-WMHypo deviations provide an individualized marker of white matter pathophysiological changes, offering novel insights for precision medicine across development, aging and brain disorders.

## Introduction

White matter (WM) abnormalities are a hallmark of brain aging and a wide spectrum of neurological disorders(*1, 2*). Among the available imaging markers, white matter hyperintensity (WMHyper) detected on T2-weighted fluid-attenuated inversion recovery (T2-FLAIR) magnetic resonance imaging (MRI) has been extensively investigated and is widely regarded as a core imaging marker of white matter injury(*3*). T2-FLAIR WMHyper (T2-WMHyper) burden has been consistently associated with cognitive decline, functional impairment, and disease progression across both vascular and neurodegenerative conditions(*4, 5*), shaping prevailing paradigms of WM pathology(*6*). However, routine clinical MRI frequently reveals additional signal alterations within WM on T1-weighted imaging (T1WI) that have received far less systematic attention. This predominant focus on T2-FLAIR-defined WM abnormalities has led to a relative neglect of other WM signal alterations that are frequently observed on routine clinical MRI.

One such feature is WM hypointensity observed on T1WI (T1-WMHypo), characterized by reduced signal intensity relative to surrounding normal-appearing WM. Previous histopathological and imaging studies have associated T1-WMHypo with heterogeneous tissue alterations, including immature WM fiber, enlarged perivascular space, severe myelin and axonal loss, cystic change or cavitation, fibrosis, mineral deposition (e.g., calcium or iron), glymphatic system abnormalities(*7–10*). Although T1-WMHypo and T2-WMHyper have often been implicitly regarded as reflecting the same underlying pathology such as WM demyelination, a few evidence indicated that these imaging features are not invariably interchangeable, as they showed distinct evolutions (increased T2-WMHyper while decreased T1-WMHypo) during the dementia progression(*11*) and some T2-WMHyper lesions didn’t show T1-hypointense in aging population(*12*). T1 WMHypo and T2 WMHyper may share both spatial and biological overlap, however, they frequently represent different stages or pathophysiological mechanisms of WM alteration(*12–14*). In particular, T1-WMHypo has been proposed to reflect more widespread or advanced tissue disruption in aging and disease conditions, such as distributed enlarged perivascular spaces in aging or severe WM damage (e.g., matrix disruption and axonal loss) in multiple sclerosis (MS), extending beyond the processes typically captured by T2-WMHyper(*7, 15–18*). Despite this potential significance, T1-WMHypo has received substantially less systematic investigation compared with T2-WMHyper detected on T2-FLAIR.

As with other structural brain measures, T1-WMHypo exhibits marked age-dependent variability across the lifespan, accompanied by substantial interindividual heterogeneity at any given age. Such variability complicates the interpretation of raw T1-WMHypo burden at the individual level and limits its clinical utility in the absence of appropriate reference standards. In this context, normative modeling has emerged as a robust framework for constructing lifespan reference curves that capture population-level variation and generate individualized deviation scores, an approach already widely applied to other brain structural metrics, including gray matter volume (GMV) and T2-WMHyper volume (T2-WMHyperV)(*19–23*). Establishing age- and sex-specific normative references for T1-WMHypo is therefore essential for contextualizing individual measurements, improving clinical interpretability, and identifying individuals with an elevated burden who may be at increased clinical risk.

In this study, leveraging a large multicenter dataset of three-dimensional T1-weighted MRI scans from 34,148 healthy volunteers, we established age- and sex-specific normative references for T1-WMHypo volume (T1-WMHypoV). We then quantified individual deviation patterns across 21 neurological and psychiatric disorders (7,587 patients) and examined the correlation between T1-WMHypoV deviations and other brain structural metrics, the value of T1-WMHypoV deviations in disease classification, clinical correlation, treatment response, and disease progression. Further, we compared the relationship and clinical relevance between T1-WMHyo and T2-WMHyper, as well as spatial-temporal analyses of T1-WMHypo across healthy controls (HCs) and brain disorders. Together, these analyses aim to assess the clinical significance of T1-WMHypo deviations independent of T2-WMHyper as an individualized imaging marker for precision medicine across development, aging and brain disorders.

## Results

### Study design

A total of 34,148 HCs and 7,587 patients with neurological and psychiatric disorders were included from 146 centers, which were drawn from the Chinese Brain Health Imaging Consortium (CHART) and the publicly available datasets including the CoRR (Consortium for Reliability and Reproducibility)(*24*), Cam-CAN (Cambridge Centre for Ageing and Neuroscience)(*25*), ABIDE (Autism Brain Imaging Data Exchange)(*26*), the ADHD-200 consortium(*27*), and OpenNeuro datasets (https://openneuro.org/). The patient cohort encompassed a broad spectrum of neurological and psychiatric conditions, with detailed diagnostic composition summarized in **Fig. 1a,b** and **Extended Data Fig. 1**.

**Fig. 1.**
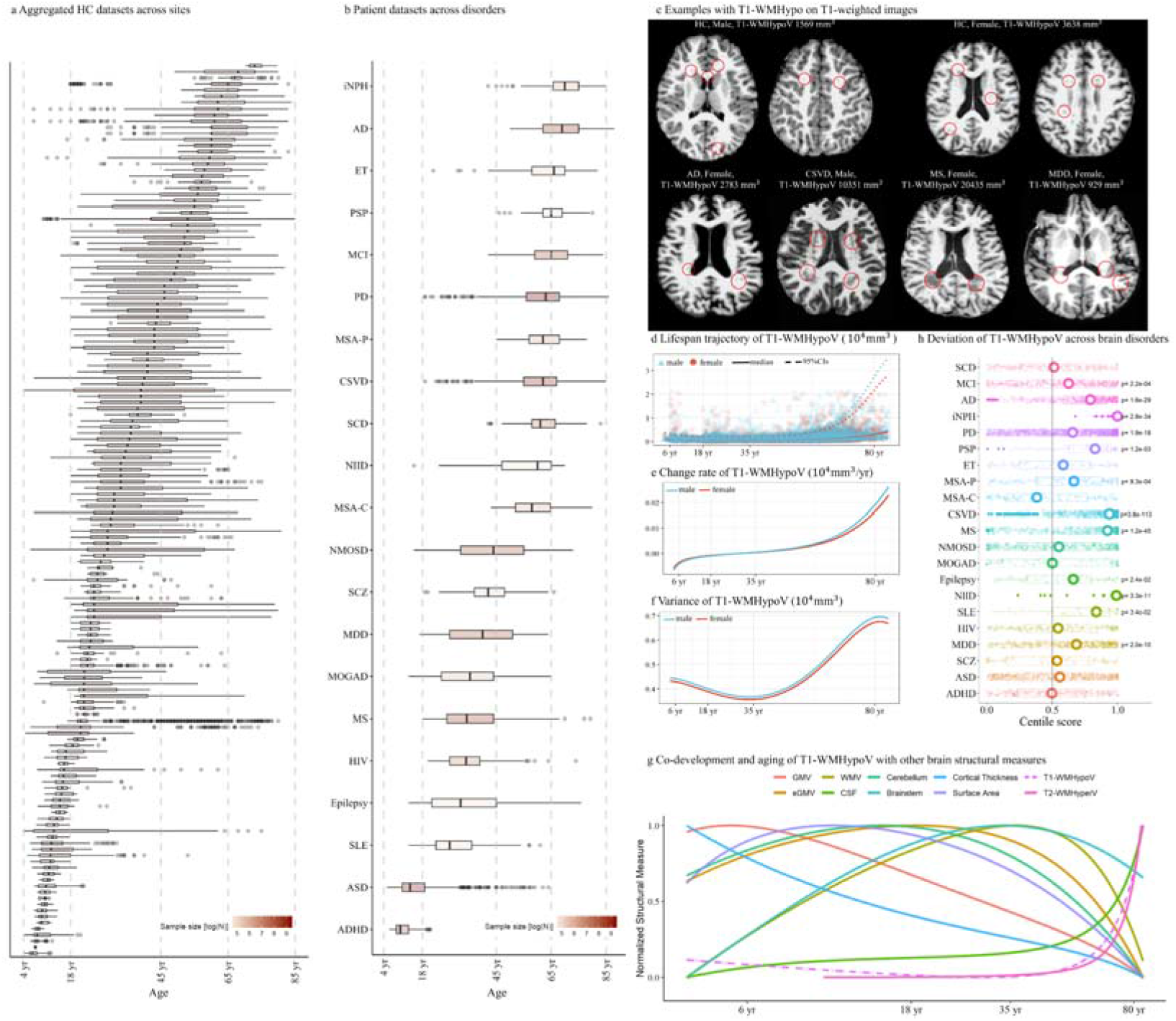
Dataset characteristics, T1-WMHypo normative reference, and deviations across 21 brain disorders. (**a**) HC data characteristics obtained from 146 international sites, the sites were arranged by the median age of the participants in each dataset; (**b**) Data characteristics of 21 brain disorders, the disorders were arranged by the median age of the participants with the disorders; (**c**) Example cases showed T1-WMHhypo on T1-wighted images across HCs and patients with brain disorders; (**d**) Normative references of T1-WMHypoV, fitted using 34,148 HCs. The curves are presented with medians and 95% confidence intervals stratified by sex; (**e**) the change rate of the T1-WMHypoV, which was calculated by the first derivative of the normative reference in (d); (**f**) The variance of the T1-WMHypoV estimated by the GAMLSS model; (**g**) Normalized curves of the T1-WMHypoV and other brain structural measures for the co-development analysis. The age was log-transformed for better visualization; (**h**) Deviations of T1-WMHypoV across 21 brain disorders including common neurological and psychiatric conditions, the presented p values were FDR-corrected. Note: T1-WMHypoV, white matter hypointensity on T1-weighted images; yr, years; HC, healthy control; SCD, subjective cognitive decline; MCI, mild cognitive impairment; AD, Alzheimer’s disease; iNPH, idiopathic normal pressure hydrocephalus; PD, Parkinson’s disease; PSP, progressive supranuclear palsy; ET, essential tremor; MSA-P, multiple system atrophy - Parkinsonian variant; MSA-C, MSA-Cerebellar variant; CSVD, cerebral small vessel disease; MS, multiple sclerosis; NMOSD, neuromyelitis optica spectrum disorders; MOGAD, myelin oligodendrocyte glycoprotein antibody-associated disease; NIID, neuronal intranuclear inclusion disease; SLE, systemic lupus erythematosus; HIV, human immunodeficiency virus; MDD, major depressive disorder; SCZ, schizophrenia; ASD, autism spectrum disorder; ADHD, attention-deficit/hyperactivity disorder; GMV, gray matter volume; sGMV, subcortical gray matter volume; WMV, white matter volume; CSF, cerebrospinal fluid; T2-WMHyperV, white matter hyperintensity on T2/FLAIR images; GAMLSS, Generalised Additive Models for Location, Scale and Shape; FDR, false discover rate.

Analyses were conducted in sequential steps. (1) Age- and sex-specific normative references of T1-WMHypoV were established using segmented T1-WMHypoV derived from three-dimensional T1WIs of HCs aged from 4 to 85 years, processed with FreeSurfer (version 7.3.2) and modeled using generalized additive models for location, scale, and shape (GAMLSS). (2) Individual T1-WMHypoV deviations (centile scores) relative to the normative reference were calculated for all participants, including both HCs and patients (for HCs, 10-fold cross-validation strategy was adopted to guarantee the individual centile scores were independent of the GMALSS model training). (3) To examine structural covariation patterns, T1-WMHypoV deviations were correlated with other brain structural metrics (normative-reference derived deviations [centile scores]), including global tissue volumes, cortical thickness, cortical surface area, and T2-WMHyperV. (4) The clinical relevance of T1-WMHypoV deviations was evaluated by assessing their added value to disease differential diagnosis, association with available clinical scores, prognostic value for treatment response and disease progression. (5) The detailed comparisons between T1-WMHypo and available T2-WMHyper were conducted. (6) Spatial-temporal analyses of regional T1-WMHypo were performed across HCs and brain disorders. Additional analyses were performed to further characterize the potential clinical relevance of T1-WMHypoV, including longitudinal analyses in participants with follow-up MR scans, and comparison analyses with absolute raw T1-WMHypoV values.

This study was conducted in accordance with the Declaration of Helsinki and complied with the Strengthening the Reporting of Observational Studies in Epidemiology (STROBE) guidelines. Ethical approval was obtained from the Ethics Committees of Beijing Tiantan Hospital, Capital Medical University (approval numbers: KY-2019-050-02, KY-2019-140-02, KY-2024-221-02, and KY-2025-203-03; Clinical Trial Number: NCT06643858). Written informed consent was obtained from all participants or their legally authorized representatives.

### Normative References of White Matter Hypointensity Volume

Lifespan normative reference of T1-WMHypoV was established using three-dimensional T1WI from 34,148 HCs aged from 4 to 85 years (median [Interquartile Range, IQR]: 37.00 [22.00, 57.00] years), including 16,711 females (48.9%) (**Table 1**). Case examples with T1-WMHypo were shown in **Fig. 1c**. The development and aging trajectory of T1-WMHypoV exhibited a J-shaped age-related pattern across the lifespan. Specifically, T1-WMHypoV showed a gradual decrease from childhood, reached its lowest point in young adulthood at 29.34 (95% confidence interval, 95%CI [28.30, 29.93]) years of age (**Fig. 1d**), and then increased progressively with advancing age. Sex-stratified analyses indicated that males exhibited slightly higher T1-WMHypoV than females across the lifespan. Inter-individual variance in T1-WMHypoV demonstrated a distinct age-dependent pattern. Variability decreased from childhood, reached a minimum at approximately 32.60 years of age, and increased thereafter, indicating relatively constrained variability in young adulthood and a widening spectrum of susceptibility to age-related T1-WMHypo changes in later life (**Fig. 1f**). The co-developmental and aging trajectories of T1-WMHypoV together with other global brain structural metrics were presented in **Fig. 1g**.

**Table 1.** Demographic and clinical information of the included healthy controls and patients with brain disorders.

|  | HC | SCD | MCI | AD | iNPH | PD | PSP | ET | MSA-P | MSA-C | CSVD | MS | NMOSD | MOGAD | Epilepsy | NIID | SLE | HIV | MDD | SCZ | ASD | ADHD |
|---|---|---|---|---|---|---|---|---|---|---|---|---|---|---|---|---|---|---|---|---|---|---|
| n | 34148 | 231 | 450 | 623 | 100 | 1351 | 53 | 61 | 113 | 106 | 904 | 612 | 443 | 115 | 55 | 46 | 53 | 173 | 439 | 62 | 1130 | 467 |
| Sex, female (NO. (%)) | 17437 (51.06) | 171 (74.03) | 286 (63.56) | 363 (58.27) | 27 (27.00) | 597 (44.19) | 16 (30.19) | 35 (57.38) | 68 (60.18) | 56 (52.83) | 382 (42.26) | 389 (63.56) | 402 (90.74) | 79 (68.70) | 28 (50.91) | 21 (45.65) | 53 (100.00) | 9 (5.20) | 283 (64.46) | 15 (24.19) | 152 (13.45) | 101 (21.63) |
| Age (yr, mean (SD)) | 39.31 (19.87) | 62.15 (6.96) | 65.15 (7.77) | 68.53 (8.51) | 69.31 (8.79) | 61.33 (9.57) | 65.13 (7.13) | 62.93 (12.23) | 62.15 (7.56) | 58.20 (8.01) | 60.67 (10.25) | 36.25 (11.95) | 43.11 (14.00) | 34.46 (13.77) | 34.69 (16.01) | 55.28 (13.64) | 30.83 (11.71) | 35.00 (9.46) | 39.36 (12.99) | 41.95 (10.00) | 15.94 (8.56) | 10.88 (3.12) |
| Education years (yr, mean (SD)) | 14.20 (4.40) | 12.68 (3.59) | 11.14 (3.89) | 10.07 (4.19) | 9.59 (3.95) | 10.81 (4.46) | 10.91 (4.18) |  | 9.72 (4.43) | 10.33 (4.18) | 8.77 (4.08) | 13.45 (3.36) | 11.28 (4.06) | 11.93 (4.32) | 12.66 (3.75) |  |  |  |  | 13.03 (3.37) |  | 6.11 (0.70) |
| MoCA (mean (SD)) | 25.24 (3.50) | 25.20 (2.96) | 20.84 (3.78) | 12.61 (6.59) | 10.51 (5.55) | 21.12 (5.14) | 15.57 (6.54) |  | 20.75 (4.84) | 19.56 (4.78) | 20.46 (5.41) | 26.20 (3.48) | 24.25 (4.42) | 24.22 (5.43) | 25.64 (4.81) |  |  |  |  |  |  |  |
| MMSE (mean (SD)) | 27.83 (2.84) | 28.37 (1.70) | 25.67 (3.13) | 17.20 (7.14) | 15.70 (6.82) | 25.66 (4.16) | 22.38 (5.59) |  | 25.86 (3.39) | 25.01 (3.86) | 24.68 (4.67) | 26.00 (6.50) | 27.72 (2.54) | 27.49 (2.98) | 28.68 (2.73) |  |  |  |  |  |  |  |
| SDMT (mean (SD)) | 52.85 (17.26) |  |  |  |  |  |  |  |  |  |  | 47.53 (15.32) | 41.93 (14.71) | 43.62 (16.12) | 53.70 (20.12) |  |  |  |  |  |  |  |
| PASAT (mean (SD)) | 44.24 (16.30) |  |  |  |  |  |  |  |  |  |  | 42.35 (10.88) | 38.78 (12.61) | 38.98 (15.83) |  |  |  |  |  |  |  |  |
| EDSS (median [IQR]) |  |  |  |  |  |  |  |  |  |  |  | 2.00 [1.00, 3.50] | 3.50 [2.00, 4.50] | 2.50 [1.88, 3.50] |  |  |  |  |  |  |  |  |
| UPDRS-III without medication (mean (SD)) |  |  |  |  |  | 49.87 (17.42) |  |  |  |  |  |  |  |  |  |  |  |  |  |  |  |  |
| HAMA (mean (SD)) | 4.80 (5.42) |  | 7.30 (6.13) | 7.89 (6.34) |  | 16.98 (8.81) |  |  |  |  |  | 4.68 (7.19) | 5.62 (6.53) |  |  |  |  |  | 18.96 (7.49) |  |  |  |
| HAMD (mean (SD)) | 4.66 (4.58) |  | 6.26 (5.52) | 7.28 (7.06) |  | 17.48 (8.89) |  |  |  |  |  | 5.41 (6.82) | 6.85 (8.70) |  |  |  |  |  | 30.04 (10.57) |  |  |  |
| PQSI (mean (SD)) | 5.29 (3.11) |  | 5.75 (3.91) | 4.89 (4.60) |  | 5.50 (2.12) |  |  |  |  |  |  |  |  |  |  |  |  |  |  |  |  |
| UPDRS-III with medication (mean (SD)) |  |  |  |  |  | 23.71 (13.39) |  |  |  |  |  |  |  |  |  |  |  |  |  |  |  |  |
| UPDRS-III without medication after DBS therapy (mean (SD)) |  |  |  |  |  | 27.56 (15.49) |  |  |  |  |  |  |  |  |  |  |  |  |  |  |  |  |
| Follow-up time (m, mean (SD)) |  |  |  |  |  |  |  |  |  |  |  | 54.57 (25.17) | 42.52 (18.27) |  |  |  |  |  |  |  |  |  |
| Follow-up EDSS (median [IQR]) |  |  |  |  |  |  |  |  |  |  |  | 2.50 [1.50, 4.00] | 3.00 [1.50, 4.75] |  |  |  |  |  |  |  |  |  |
| EDSS Progression (%) |  |  |  |  |  |  |  |  |  |  |  | 80 (34.19) | 40 (31.75) |  |  |  |  |  |  |  |  |  |
| SPMS conversion (%) |  |  |  |  |  |  |  |  |  |  |  | 44 (18.72) |  |  |  |  |  |  |  |  |  |  |

Note: n, number; yr, years; m, months, SD, standard deviation; IQR, interquartile range; DBS, deep brain stimulation; HC, healthy control; SCD, subjective cognitive decline; MCI, mild cognitive impairment; AD, Alzheimer’s disease; iNPH, idiopathic normal pressure hydrocephalus; PD, Parkinson’s disease; PSP, progressive supranuclear palsy; ET, essential tremor; MSA-P, multiple system atrophy - Parkinsonian variant; MSA-C, MSA-Cerebellar variant; CSVD, cerebral small vessel disease; MS, multiple sclerosis; NMOSD, neuromyelitis optica spectrum disorders; MOGAD, myelin oligodendrocyte glycoprotein antibody-associated disease; NIID, neuronal intranuclear inclusion disease; SLE, systemic lupus erythematosus; HIV, human immunodeficiency virus; MDD, major depressive disorder; SCZ, schizophrenia; ASD, autism spectrum disorder; ADHD, attention-deficit/hyperactivity disorder; GMV, gray matter volume; sGMV, subcortical gray matter volume; WMV, white matter volume; CSF, cerebrospinal fluid; TIV, total intracranial volume; T2-WMHyperV, white matter hyperintensity on T2/FLAIR images; MoCA, Montreal Cognitive Assessment; MMSE, Mini-Mental State Examination; SDMT, Symbol Digit Modalities Test; PASAT, Paced Auditory Serial Addition Test; EDSS, Expanded Disability Status Scale; UPDRS-III, Unified Parkinson’s Disease Rating Scale - Part III; HAMA, Hamilton Anxiety Rating Scale; HAMD, Hamilton Depression Rating Scale; PQSI, Pittsburgh Sleep Quality Index; SPMS, secondary progressive multiple sclerosis. Categorical data are presented as percentages. Continuous data are presented as the means and standard deviations. Ranked data are displayed as medians and interquartile ranges.

### White Matter Hypointensity Deviations Across Neurological and Psychiatric Disorders

Individualized T1-WMHypoV deviation scores were calculated for HCs and patients using the normative reference framework. Group differences between patients and age- and sex-matched HCs, with additional site matching when available, were evaluated using Monte Carlo permutation tests. Effect sizes were quantified using Cohen’s d, and statistical significance p values were determined after false discovery rate (FDR) correction across disorders (**Fig. 1h**). Negative effect sizes indicated higher T1-WMHypo burden in patients relative to HCs.

Significant T1-WMHypoV deviations were observed in the majority of neurological disorders (11/17, 65%) and only major depressive disorder (MDD) in psychiatric disorders (1/4, 25%), with marked heterogeneity in effect magnitude. Large deviations (|d| ≥ 0.8) were detected in idiopathic normal pressure hydrocephalus (iNPH, Cohen’s d = -2.25, 95%CI [-2.62, -1.88], pFDR < 0.001), neuronal intranuclear inclusion disease (NIID, d = -1.65 [-2.13, -1.17], pFDR < 0.001), cerebral small vessel disease (CSVD, d = -1.26 [-1.37, -1.15], pFDR < 0.001), and MS (d = -0.85 [-1.09, -0.74], pFDR < 0.001). Moderate deviations (0.5 ≤ |d| < 0.8) were observed in Alzheimer’s disease (AD, d = -0.74 [-0.87, -0.61], pFDR < 0.001) and progressive supranuclear palsy (PSP, d = -0.69 [-1.08, -0.30], pFDR < 0.001). Small yet statistically significant deviations (|d| ≥ 0.2) were detected in epilepsy (d = -0.48 [-0.86, -0.10], pFDR = 0.013), multiple system atrophy-Parkinsonian variant (MSA-P, d = -0.48 [-0.74, -0.21], pFDR < 0.001), systemic lupus erythematosus (SLE, d = -0.46 [-0.84, -0.07], pFDR < 0.020), MDD (d = -0.45 [-0.58, -0.31], pFDR < 0.001), Parkinson’s disease (PD, d = -0.35 [-0.43, -0.27], pFDR < 0.001) and mild cognitive impairment (MCI, d = -0.28 [-0.43, -0.14], pFDR < 0.001).

In contrast, no significant T1-WMHypoV deviations were observed in subjective cognitive decline (SCD), essential tremor (ET), MSA-Cerebellar variant (MSA-C), neuromyelitis optica spectrum disorders (NMOSD), myelin oligodendrocyte glycoprotein antibody-associated disease (MOGAD), human immunodeficiency virus (HIV), schizophrenia (SCZ), autism spectrum disorder (ASD), or attention-deficit/hyperactivity disorder (ADHD).

Pairwise comparisons across disease groups (**Extended Data Fig. 2**) further highlighted disorder-specific differences in T1-WMHypoV deviation patterns. Together, these results demonstrate that T1-WMHypoV burden varies substantially across neurological and psychiatric conditions, reflecting heterogeneous manifestations of white matter abnormality.

### Correlations between White Matter Hypointensity and Other Brain Structural Metrics

Associations of normative reference-derived deviations between T1-WMHypoV and other brain structural metrics were examined across HCs and all disease groups (**Fig. 2a)**. HCs were stratified into younger (≤29 years), middle-aged (29-57 years), and older (≥57 years) subgroups according to key inflection points of the T1-WMHypoV lifespan trajectory (**Fig. 1d**). Primary results are reported using partial Pearson correlations adjusted for age, sex, and total intracranial volume (TIV), with univariate Pearson correlations and additional adjustment for T2-WMHyperV performed for comparison. Here, correlations with FDR-corrected p < 0.05 were considered statistically significant and reported, while FDR-uncorrected results are shown for exploratory purpose.

**Fig. 2.**
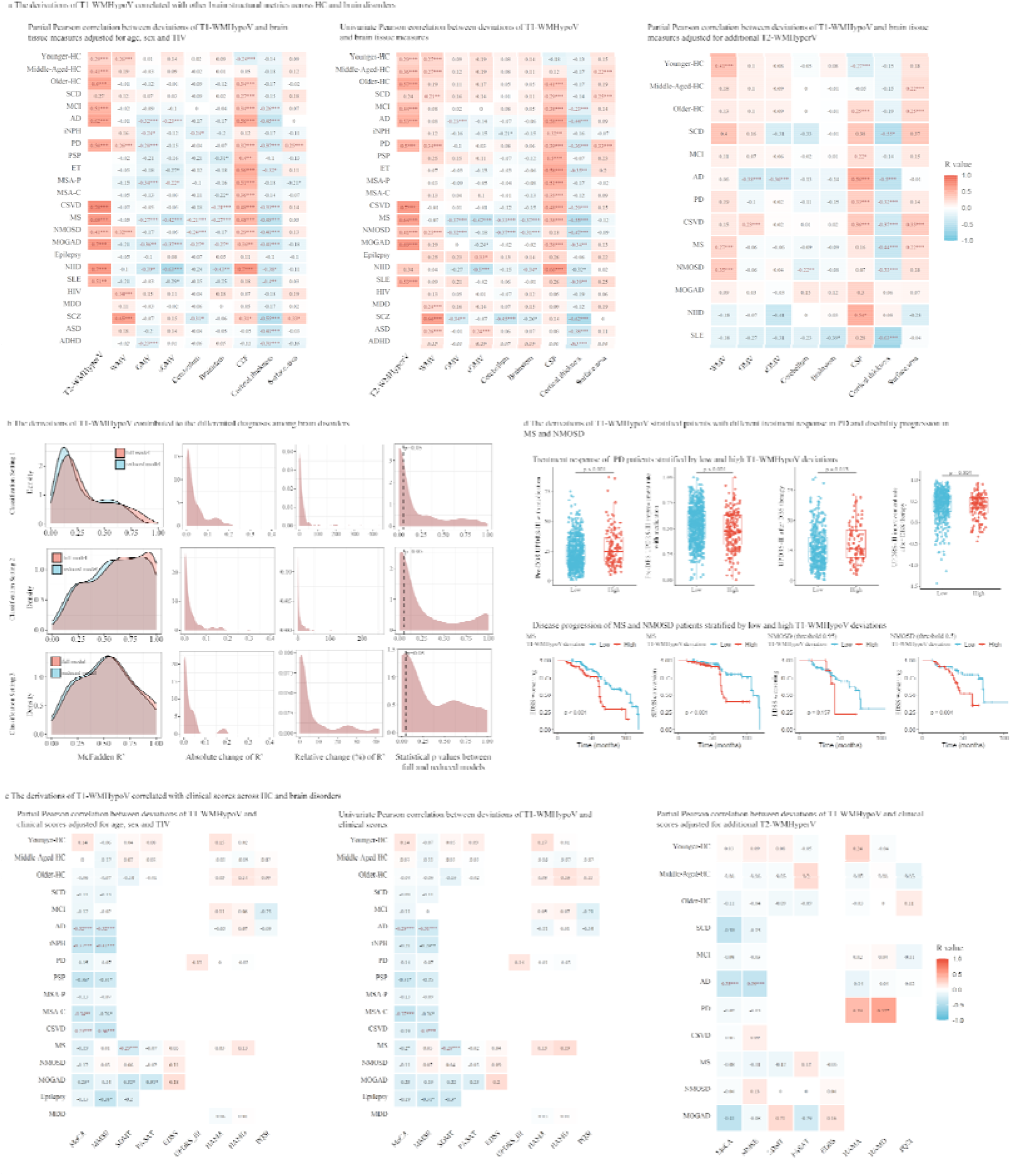
Structural correlation and clinical relevance of T1-WMHypoV deviations. (**a**) Correlation of centile scores between T1-WMHypoV and other brain structural metrics using partial Pearson correlations adjusted for age, sex, and TIV, univariate correlations and partial Pearson correlations additionally adjusted for T2-WMHyperV. Correlations with FDR-corrected p < 0.05 were considered significant (labelled in red and with *), while uncorrected results are shown for exploratory purposes (only labeled with *); (**b**) The T1-WMHypo contributes to disease classification across neurological and psychiatric disorders. The distribution of the McFadden R²of both full and reduced models, the absolute (defined as [McFadden R² of full model - McFadden R² of reduced model]) and relative (defined as [McFadden R² of full model - McFadden R² of reduced model]/McFadden R² of reduced model*100%) changes of McFadden R², and the p values distribution across the paired classifications among brain disorders were presented; (**c**) Associations between T1-WMHypoV centile scores and clinical scores using partial Pearson correlations adjusted for age, sex, and TIV, univariate correlations and partial Pearson correlations additionally adjusted for T2-WMHyperV; (**d**) Treatment response and disease progression assessment by T1-WMHypoV deviations in PD, MS and NMOSD. Treatment response assessment in PD by comparing the UPDRS-III prior to and after medication or DBS therapy. Here, we defined the PD cases with extreme deviations (centile score > 0.95) as the high T1-WMHypo burden subgroup, and the remaining as the low T1-WMHypo burden subgroup. Between-group differences on motor improvement after medication and one-year follow-up after deep brain stimulation (DBS) therapy were analyzed with Student’s t-test. P value was uncorrected; Above deviation cutoffs were also applied for MS and NMOSD cases to define the high and low T1-WMHypo burden subgroups. Kaplan-Meier analysis with a log-rank test was conducted to investigate the potential differences in disease progression of the stratified subgroups. P value was uncorrected. Note: T1-WMHypoV, white matter hypointensity on T1-weighted images; AUC, area under the curve; Acc, accuracy; IQR, Interquartile Range; HC, healthy control; SCD, subjective cognitive decline; MCI, mild cognitive impairment; AD, Alzheimer’s disease; iNPH, idiopathic normal pressure hydrocephalus; PD, Parkinson’s disease; PSP, progressive supranuclear palsy; ET, essential tremor; MSA-P, multiple system atrophy - Parkinsonian variant; MSA-C, MSA-Cerebellar variant; CSVD, cerebral small vessel disease; MS, multiple sclerosis; NMOSD, neuromyelitis optica spectrum disorders; MOGAD, myelin oligodendrocyte glycoprotein antibody-associated disease; NIID, neuronal intranuclear inclusion disease; SLE, systemic lupus erythematosus; HIV, human immunodeficiency virus; MDD, major depressive disorder; SCZ, schizophrenia; ASD, autism spectrum disorder; ADHD, attention-deficit/hyperactivity disorder; GMV, gray matter volume; sGMV, subcortical gray matter volume; WMV, white matter volume; CSF, cerebrospinal fluid; TIV, total intracranial volume; T2-WMHyperV, white matter hyperintensity on T2-FLAIR images; MoCA, Montreal Cognitive Assessment; MMSE, Mini-Mental State Examination; SDMT, Symbol Digit Modalities Test; PASAT, Paced Auditory Serial Addition Test; EDSS, Expanded Disability Status Scale; UPDRS-III, Unified Parkinson’s Disease Rating Scale - Part III; HAMA, Hamilton Anxiety Rating Scale; HAMD, Hamilton Depression Rating Scale; PQSI, Pittsburgh Sleep Quality Index; DBS, deep brain stimulation; SPMS, Secondary Progressive MS; FDR, false discovery rate; Classification Setting 1 (210 disease pairs): logistic regression between each pair of diseases by variates including brain structural metrics (WMV, GMV, sGMV, cerebellum, brainstem, CSF, cortical thickness, surface area and TIV) and T1-WMHypoV (full model), versus above metrics excluding T1-WMHypoV (reduced model); Classification Setting 2 (210 disease pairs): logistic regression between each pair of diseases by variates including brain structural metrics (WMV, GMV, sGMV, cerebellum, brainstem, CSF, cortical thickness, surface area and TIV), demographics (age and sex) and T1-WMHypoV (full model), versus above metrics excluding T1-WMHypoV (reduced model); Classification Setting 3 (45 disease pairs): logistic regression between each pair of diseases by variates including brain structural metrics (WMV, GMV, sGMV, cerebellum, brainstem, CSF, cortical thickness, surface area, TIV and additional T2-WMHyperV), demographics (age and sex) and T1-WMHypoV (full model), versus above metrics excluding T1-WMHypoV (reduced model). * p< 0.05, ** p< 0.005, *** p< 0.001.

Across diagnostic groups, T1-WMHypoV showed positive correlations with T2-WMHyperV in all available disease groups except SCD (r = 0.29 to 0.78). Positive associations with white matter volume (WMV) were observed in younger HCs and in PD, NMOSD, HIV, and SCZ (r = 0.26 to 0.65). Associations with cerebrospinal fluid (CSF) volume displayed an age-dependent pattern in HCs, with negative correlations in younger individuals (r = -0.24) and positive correlations in older individuals (r = 0.34), whereas positive correlations were present in most (14/21) disorders (r = 0.27 to 0.70), except iNPH, epilepsy, SLE, HIV, MDD, ASD, and ADHD. Positive associations with cortical surface area were detected in PD and SCZ (r = 0.25 to 0.33).

In contrast, T1-WMHypoV showed consistent negative correlations with cortical GMV in AD, iNPH, PD, MSA-P, MS, MOGAD, NIID, and ADHD (r = -0.39 to -0.23), and with subcortical GMV (sGMV) in AD, MS, MOGAD, and NIID (r = -0.63 to -0.22). Additional negative associations were observed with cerebellar volume in iNPH, MS, NMOSD, MOGAD, and SCZ, and with brainstem volume in CSVD, MS, MOGAD, and NIID. Mean cortical thickness was negatively correlated with T1-WMHypoV across most (13/21) disorders, including MCI, AD, PD, ET, CSVD, MS, NMOSD, MOGAD, NIID, SLE, SCZ, ASD, and ADHD.

Univariate analyses and partial correlations additionally adjusted for T2-WMHyperV yielded largely consistent correlations, with attenuated effect sizes (r values) after accounting for conventional T2-WMHyper burden.

### Clinical Relevance of White Matter Hypointensity on Differential Diagnoses, Clinical Associations and Disease Outcomes

#### White Matter Hypointensity Contributed to Disease Differential Diagnoses

To assess whether the T1-WMHypoV has additional contribution to the disease differential diagnosis, we performed a series of classifications among the above brain disorders (HCs were not considered in such analyses, as we focused on the disease differential diagnoses) using logistic regression. To quantify the potential contribution of T1-WMHypoV deviations, we compared the adjusted McFadden R² between full (all variates were considered) and reduced models (all variates except T1-WMHypoV were considered) for each pair of disorder groups by following three classification settings (**Fig. 2b**). Setting 1 (210 disease pairs): logistic regression between each pair of diseases by variates including brain structural metrics (WMV, GMV, sGMV, cerebellum, brainstem, CSF, cortical thickness, surface area and TIV) and T1-WMHypoV (full model), versus above metrics excluding T1-WMHypoV (reduced model); Setting 2 (210 disease pairs): logistic regression between each pair of diseases by variates including brain structural metrics (WMV, GMV, sGMV, cerebellum, brainstem, CSF, cortical thickness, surface area and TIV), demographics (age and sex) and T1-WMHypoV (full model), versus above metrics excluding T1-WMHypoV (reduced model); and Setting 3 (45 disease pairs due to that the T2-FLAIR was only available in ten disease groups) logistic regression between each pair of diseases by variates including brain structural metrics (WMV, GMV, sGMV, cerebellum, brainstem, CSF, cortical thickness, surface area, TIV and additional T2-WMHyperV), demographics (age and sex) and T1-WMHypoV (full model), versus above metrics excluding T1-WMHypoV (reduced model). Likelihood Ratio Test was adopted to test the model performances between each paired full and reduced models, a p < 0.05 was considered statistically significant. Here, we reported the McFadden R²of both full and reduced models, the absolute (defined as [McFadden R² of full model - McFadden R² of reduced model]) and relative (defined as [McFadden R² of full model - McFadden R² of reduced model]/McFadden R² of reduced model*100%) changes of McFadden R², and the p values distribution across the paired classifications among brain disorders.

As shown in **Fig. 2b**, we observed the T1-WMHypoV significantly contributed (that is the full model performance (McFadden R²) was improved compared to reduced model by Likelihood Ratio Test with p < 0.05) to classifications of 123/210 (58.57%), 102/210 (48.57%), 19/45 (42.22%) disease pairs for classification setting 1-3, respectively. The added McFadden R²by T1-WMHypoV were 0.018 [0.00, 0.21] (median [range]), 0.0090 [0.00, 0.30], and 0.010 [0.00, 0.19] for classification setting 1-3, respectively. Relative increments of McFadden R² by T1-WMHypoV were 8.65% [0.00%, 434.90%], 1.66% [0.00%, 254.47%], and 1.19% [0.00%, 41.83%] for classification setting 1-3, respectively.

#### White Matter Hypointensity Associated with Clinical Scores

Associations between T1-WMHypoV centile scores and clinical scores (cognitive, motor, mood and sleep scores, see details in **Table 1**) were examined across HC and disease groups (**Fig. 2c**). Here, we stratified the HCs into three subgroups by age as above. Similar Pearson correlation analyses were conducted as above. An FDR-corrected p < 0.05 across all diagnostic groups and measures was considered statistically significant. Uncorrected statistically significant findings were also reported to provide hints of the potential clinical relevance of T1-WMHypoV.

After adjustment for age, sex and TIV, higher T1-WMHypoV was consistently associated with poorer global cognitive performance across multiple disorders. Significant negative correlations with Montreal Cognitive Assessment (MoCA, r = -0.23 to -0.37) were observed in AD, iNPH, MSA-C and CSVD, as well as PSP (FDR-unsurvived) and MOGAD (FDR-unsurvived). Similar negative associations with Mini-Mental State Examination (MMSE, r = -0.26 to -0.41) were observed in AD, iNPH, and CSVD, as well as PSP (FDR-unsurvived) and MSA-C (FDR-unsurvived). In addition, negative association with MMSE (r = -0.38) was observed in epilepsy (FDR-unsurvived)

In MS and MOGAD (FDR-uncorrected), higher T1-WMHypoV was associated with lower Symbol Digit Modalities Test (SDMT) performance (r = -0.29 and -0.32, respectively). In addition, T1-WMHypoV showed a negative correlation with Paced Auditory Serial Addition Test (PASAT) scores in MOGAD (r = -0.31, FDR-unsurvived). No associations were observed between T1-WMHypoV and Expanded Disability Status Scale (EDSS) across neuroinflammatory disorders.

Motor-, mood- and sleep-related scores showed largely no significant associations with T1-WMHypoV across disorders. Univariate analyses and partial correlations with additional adjustment for T2-WMHyperV yielded comparable findings.

#### White Matter Hypointensity Stratified Patients with Different Treatment Response and Disease Progression

We further examined the potential predictive ability of T1-WMHypoV for treatment response in PD, and disease progression in MS and NMOSD. For treatment response assessment in PD, we defined the PD cases with extreme deviations (centile score > 0.95) as the high T1-WMHypo burden subgroup, and the remaining as the low T1-WMHypo burden subgroup. Between-group differences in motor improvement after medication and one-year follow-up after deep brain stimulation (DBS) therapy were analyzed by Student’s t-test. The same deviation cutoffs were also applied for MS and NMOSD cases to define the high and low T1-WMHypo burden subgroups. Kaplan-Meier analysis with a log-rank test was conducted to investigate the potential differences in disease progression (EDSS worsening in MS and NMOSD, and conversion to secondary progressive MS [SPMS] from relapsing-remitting MS [RRMS]) of the stratified subgroups. We additionally conducted multivariate Cox proportional hazards regression to test the independent predictive value of T1-WMHypoV deviations, by considering age, sex, and other brain structural metrics.

In PD, patients with high T1-WMHypoV deviations showed more severe motor impairment than those with low deviations, reflected by higher Unified Parkinson’s Disease Rating Scale-Part III (UPDRS-III) scores both under medication and in the DBS-off state (both p < 0.001, **Fig. 2d**). Patients in the high T1-WMHypoV subgroup demonstrated lower motor improvement in response to medication compared with those in the low T1-WMHypoV subgroup (p = 0.013). In contrast, no significant difference in motor improvement following DBS was observed between subgroups (p = 0.304), indicating that T1-WMHypoV burden is associated with pharmacological responsiveness but does not predict DBS-related benefit.

In MS, patients with high T1-WMHypoV deviations exhibited a worse disease course than those with low deviations (**Fig. 2d**). Kaplan-Meier analyses demonstrated that the patients with high-deviation subgroup showed a greater risk of EDSS worsening (p < 0.001) and an increased risk of conversion from relapsing-remitting multiple sclerosis (RRMS, shortened as MS in the above contents) to SPMS (p < 0.001). At 5 years of follow-up, the cumulative probability of EDSS worsening was 38.91% [28.80%, 47.59%] in the high-deviation group compared with 22.73% [95%CI, 9.15%, 34.29%] in the low T1-WMHypoV group. The cumulative probability of SPMS conversion was 24.22% [14.37%, 32.95%] in the high-deviation group compared with 9.93% [0.00%, 18.89%] in the low T1-WMHypoV group. In NMOSD, stratification using the high-deviation threshold (centile > 0.95) did not reveal a significant difference in disability progression between subgroups (p = 0.157). However, when a lower threshold (centile > 0.50) was applied, patients with high T1-WMHypoV deviations showed a greater risk of EDSS worsening compared with those with low deviations (p = 0.004). At 5 years of follow-up, the cumulative probability of EDSS worsening was 65.08% [38.83%, 80.07%] in the high-deviation group compared with 20.39% [7.06%, 31.80%] in the low T1-WMHypoV group.

Across demyelinating disorders, the prognostic relevance of T1-WMHypoV deviations was further supported by multivariate Cox regression analyses, in which higher T1-WMHypoV deviations remained associated with increased risk of disability progression after adjustment for demographic factors, and other brain structural metrics in MS (**Extended Data Fig. 3**).

### Comparison of Normative Reference and Clinical Relevance with T2-WMHyperV across HCs and Brain Disorders

Based on the available T2-WMHyperV (n=2,276 for HCs, and n=2,939 for brain disorders), we first analyzed the correlations between absolute raw T1-WMHyoV and T2-WMHyperV by univariate Pearson correlation across HC and disease groups.

Then we fitted the normative reference of T2-WMHyperV with the same method as described in the T1-WMHypoV normative reference construction. The deviations of T2-WMHyperV across HCs and brain disorders were calculated and compared as described in the T1-WMHypoV deviation analysis. Univariate Pearson correlation with other brain structural metrics and clinical scores was conducted to test the clinical relevance of T2-WMHyperV deviations for comparison with those in T1-WMHypoV. Comparisons between T1-WMHypoV and T2-WMHyperV were performed in different conditions (in all cases and subsets with low T2-WMHyperV [volume < 1ml]).

We first analyzed the relationship between raw T1-WMHypoV and T2-WMHyperV across HC lifespan and brain disorders (**Fig. 3a** and **b**). The example cases showed that T1-WMHyo on T1WI and T2-WMHyper on T2-FLAIR have both spatially overlapping and nonoverlapping distributions in normal development and aging as well as across brain disorders. Further, we found significant correlations (r = 0.31 to 0.80) between raw T1-WMHypoV and T2-WMHyperV across HC and brain disorder groups except for SCD. In HC subgroups, we observed the correlations (r values) increased from 0.31 to 0.69 in Younger, Middle-aged and Older HC subgroups. In brain disorders, the correlation was highest (r = 0.80, pFDR < 0.001) in CSVD and lowest in SCD (r = 0.37, pFDR = 0.078), indicating the T1-WMHypo and T2-WMHyper may share more underlying pathology especially in conditions with potential high cerebral vascular risks.

**Fig. 3.**
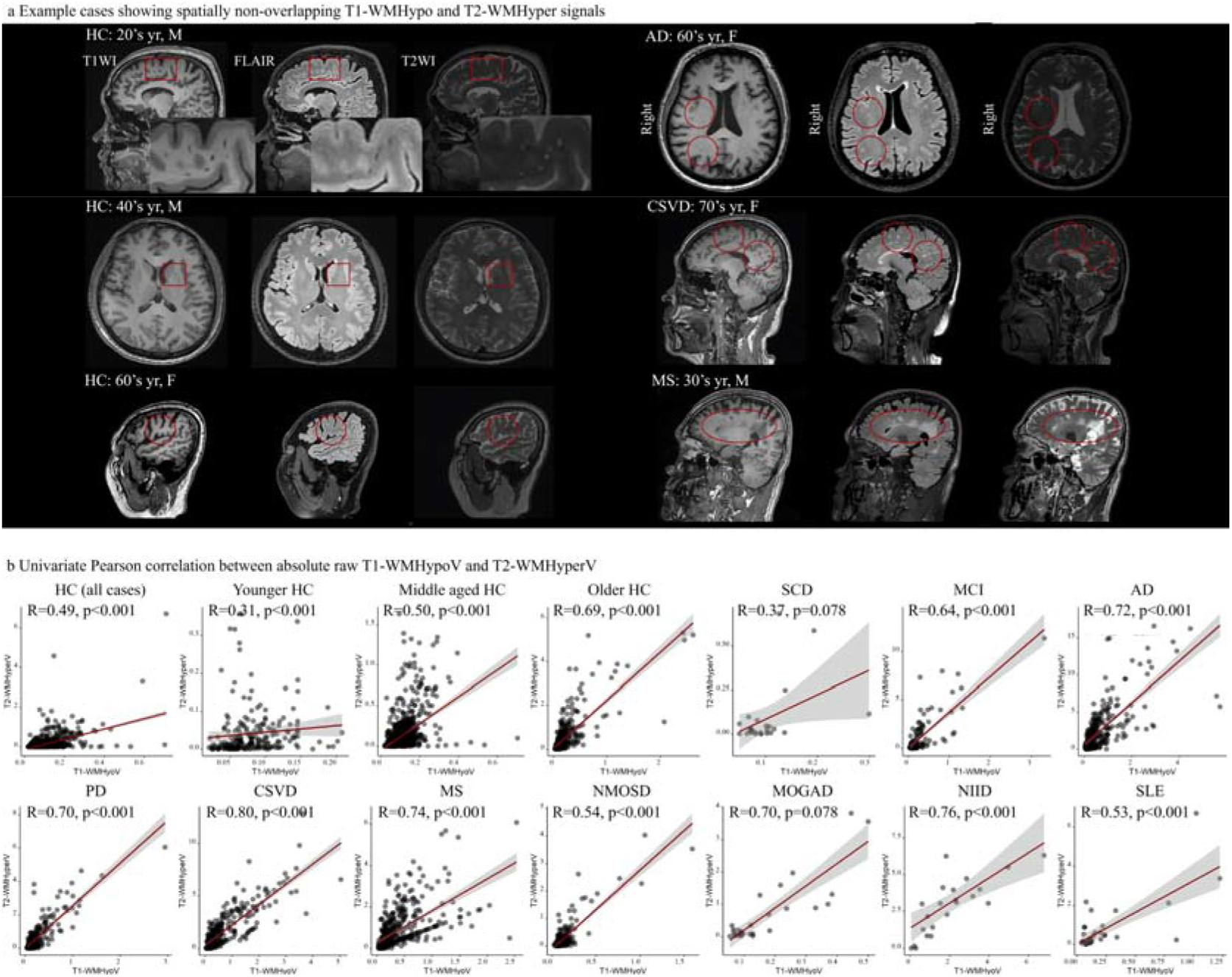
Relationship between T1-WMHypo on T1WI and T2-WMHyper on T2-FLAIR. (**a**) Example cases that showed spatially overlapping and nonoverlapping T1-WMHypo on T1WI and T2-WMHyperV on FLAIR; (**b**) Univariate Pearson correlation of absolute raw T1-WMHypoV and T2-WMHyperV. Correlations with FDR-corrected p < 0.05 were considered statistically significant. Note: T1-WMHypoV, white matter hypointensity on T1-weighted images; T2-WMHyperV, white matter hyperintensity on T2-FLAIR images; yr, years; HC, healthy control; AD, Alzheimer’s disease; CSVD, cerebral small vessel disease; MS, multiple sclerosis; MDD, major depressive disorder; FDR, false discovery rate.

The intersection age of T1-WMHypoV and T2-WMHyperV was observed at about 57 years (**Fig. 4a**). Before this intersection age, the T1-WMHypoV was predominant compared to T2-WMHyperV, and after this, T2-WMHyperV increased rapidly and became predominant compared to T1-WMHypoV. We observed similar T2-WMHyperV deviations as those in T1-WMHypoV in most of the brain disorders, in contrast, the NMOSD and MOGAD showed significant higher T2-WMHyperV deviations, which showed no T1-WMHypoV deviations (**Fig. 4b**). We also observed that the T2-WMHyperV correlated with other brain structural metrics (negative r ranged from -0.65 to -0.20, and positive r ranged from 0.23 to 0.51, all pFDR < 0.05), similar to those findings in T1-WMHypoV across HCs and brain disorders (**Fig. 4c**). Similar correlations with clinical scores were observed (negative r ranges from -0.47 to 0.22, positive correlation with MoCA in Younger-HC [r=0.30], all pFDR < 0.05) (**Fig. 4d**) between T2-WMHyperV and T1-WMHypoV. For those cases with low T2-WMHyperV (volume < 1ml), the T1-WMHypo showed significant larger volume than T2-WMHyper (**Fig. 4f**), indicating the predominant pathologies reflected by T1-WMHypo at low T2-WMHyper burden. Meanwhile in the low T2-WMHyperV cases, the T1-WMHypoV deviations largely preserved the correlations with other brain structural metrics and clinical scores (**Fig. 4g, h**), which implied T1-WMHypo is a sensitive and clinically relevant marker of WM alterations at low or absent burden of T2-WMHyper.

**Fig. 4.**
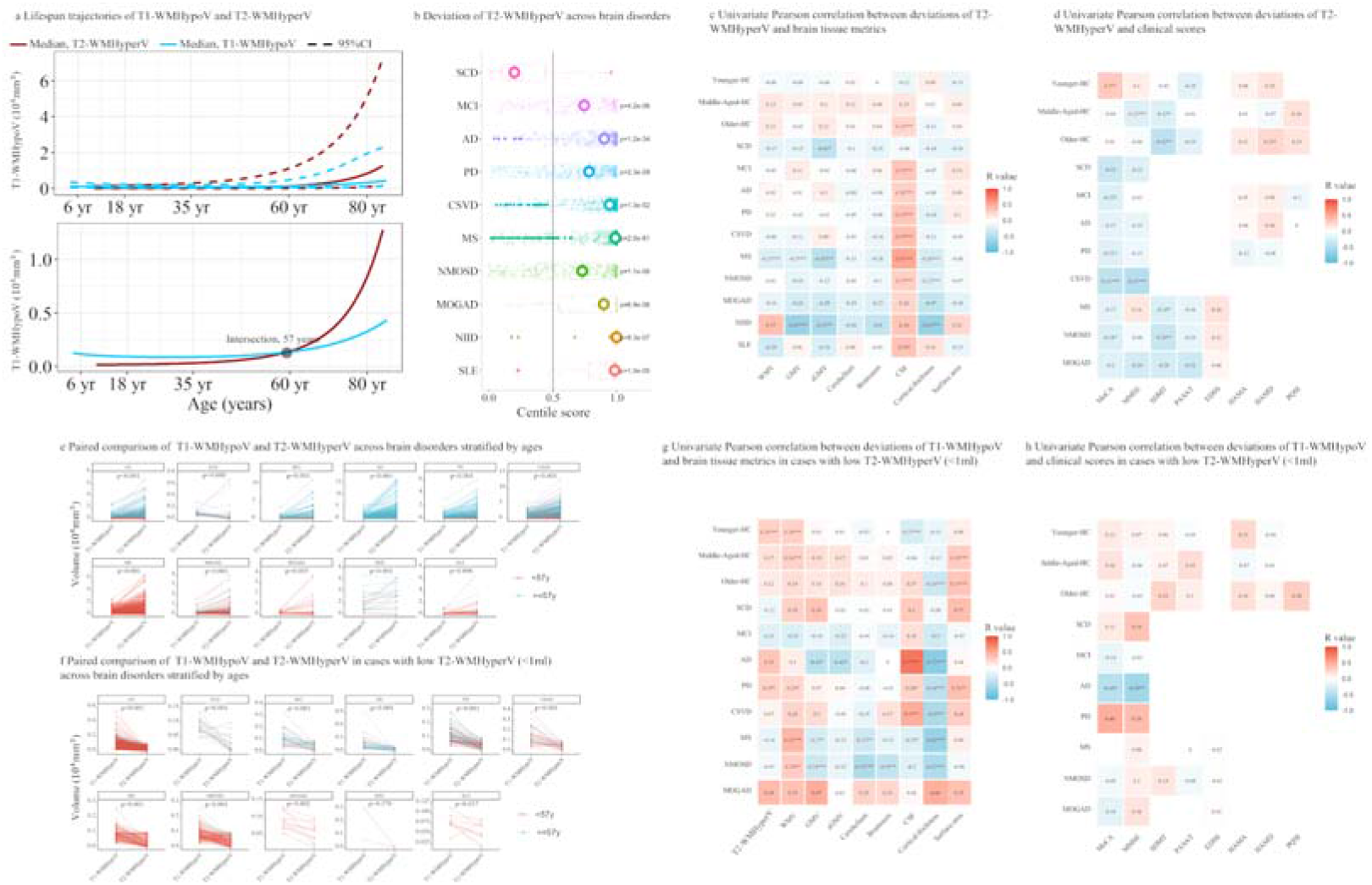
Comparison analyses of lifespan trajectory and clinical relevance between T1-WMHypoV and T2-WMHyperV. (**a**) Lifespan trajectories of T1-WMHypoV and T2-WMHyperV; (**b**) Deviations of T2-WMHyperV across available brain disorders, the presented p values were FDR-corrected; (**c**) Univariate Pearson correlation of T2-WMHyperV with other brain structural metrics. Correlations with FDR-corrected p < 0.05 were considered significant (labelled in red and with *), while uncorrected results are shown for exploratory purposes (only labeled with *); (**d**) Univariate Pearson correlation of T2-WMHyperV with clinical scores; (**e**) Comparison of T1-WMHyoV and T2-WMHyperV across available groups; (**f**) Comparison of T1-WMHyoV and T2-WMHyperV across available cases with low T2-WMHyperV (volume < 1ml); (**g**) Univariate Pearson correlation of T1-WMHyoV with other brain structural metrics in cases with low T2-WMHyperV; (**h**) Univariate Pearson correlation of T2-WMHyperV with clinical scores in cases with low T2-WMHyperV. Note: T1-WMHypoV, white matter hypointensity on T1-weighted images; yr, years; HC, healthy control; SCD, subjective cognitive decline; MCI, mild cognitive impairment; AD, Alzheimer’s disease; iNPH, idiopathic normal pressure hydrocephalus; PD, Parkinson’s disease; CSVD, cerebral small vessel disease; MS, multiple sclerosis; NMOSD, neuromyelitis optica spectrum disorders; MOGAD, myelin oligodendrocyte glycoprotein antibody-associated disease; NIID, neuronal intranuclear inclusion disease; SLE, systemic lupus erythematosus; HIV, human immunodeficiency virus; GMV, gray matter volume; sGMV, subcortical gray matter volume; WMV, white matter volume; CSF, cerebrospinal fluid; T2-WMHyperV, white matter hyperintensity on T2-FLAIR images; MoCA, Montreal Cognitive Assessment; MMSE, Mini-Mental State Examination; SDMT, Symbol Digit Modalities Test; PASAT, Paced Auditory Serial Addition Test; EDSS, Expanded Disability Status Scale; HAMA, Hamilton Anxiety Rating Scale; HAMD, Hamilton Depression Rating Scale; PQSI, Pittsburgh Sleep Quality Index; FDR, false discovery rate. * p< 0.05, ** p< 0.005, *** p< 0.001.

### Spatial-Temporal Patterns of White Matter Hypointensity across HCs and Brain Disorders

To further reveal the regional characteristics of T1-WMHypo, we first normalized segmented T1-WMHypo into Montreal Neurological Institute (MNI) space across different HC subgroups (16 subgroups spanning ages 4 to 85 years, at 5-year intervals) and brain disorders, and the spatial probability maps of T1-WMHypo distribution were obtained for visualization. Then, we defined the regional T1-WMHypoV according to the integrated parcellations based on the Hammers atlas(*28, 29*). Seven regional T1-WMHypoV metrics, including averaged T1-WMHypoVs within corpus callosum, basal ganglia, frontal, parietal, temporal, occipital and insula regions, were calculated. The normative references of these regional T1-WMHypoV were constructed by GAMLSS as described above, and the regional T1-WMHypoV deviations across brain disorders were obtained. Statistical analyses on these regional deviations were the same as above.

First, we presented the spatial analyses of the T1-WMHypoV across HCs and brain disorders (**Fig. 5**). The spatial T1-WMHypo distribution showed a consistent temporal (along the age axis) pattern with the development and aging patterns of whole brain T1-WMHypoV. Specifically, the T1-WMHypo distribution involved the periventricular regions as well as gray-white matter junction especially in children aged 4-10 years, and then the distribution of gray-white matter junction disappeared, and was mainly located around periventricular regions during the younger and middle life, and then started to expand from the periventricular regions to deep WM with the aging process. The development and aging trajectories of these regional T1-WMHypoV were consistent with those of whole brain T1-WMHypoV, showing J-shaped trajectories, reaching the lowest burden around 25 (median, 24.84 and range, 21.77-28.40) years. In addition, disorder-specific spatial deviations of T1-WMHypoV were observed, indicating the different brain regional involvements across disorders. These regional T1-WMHypoV normative references and their derived deviations provided additional information for the interpretation of pathophysiology in normal development and aging, and pathology across both neurological and psychiatric disorders.

**Fig. 5.**
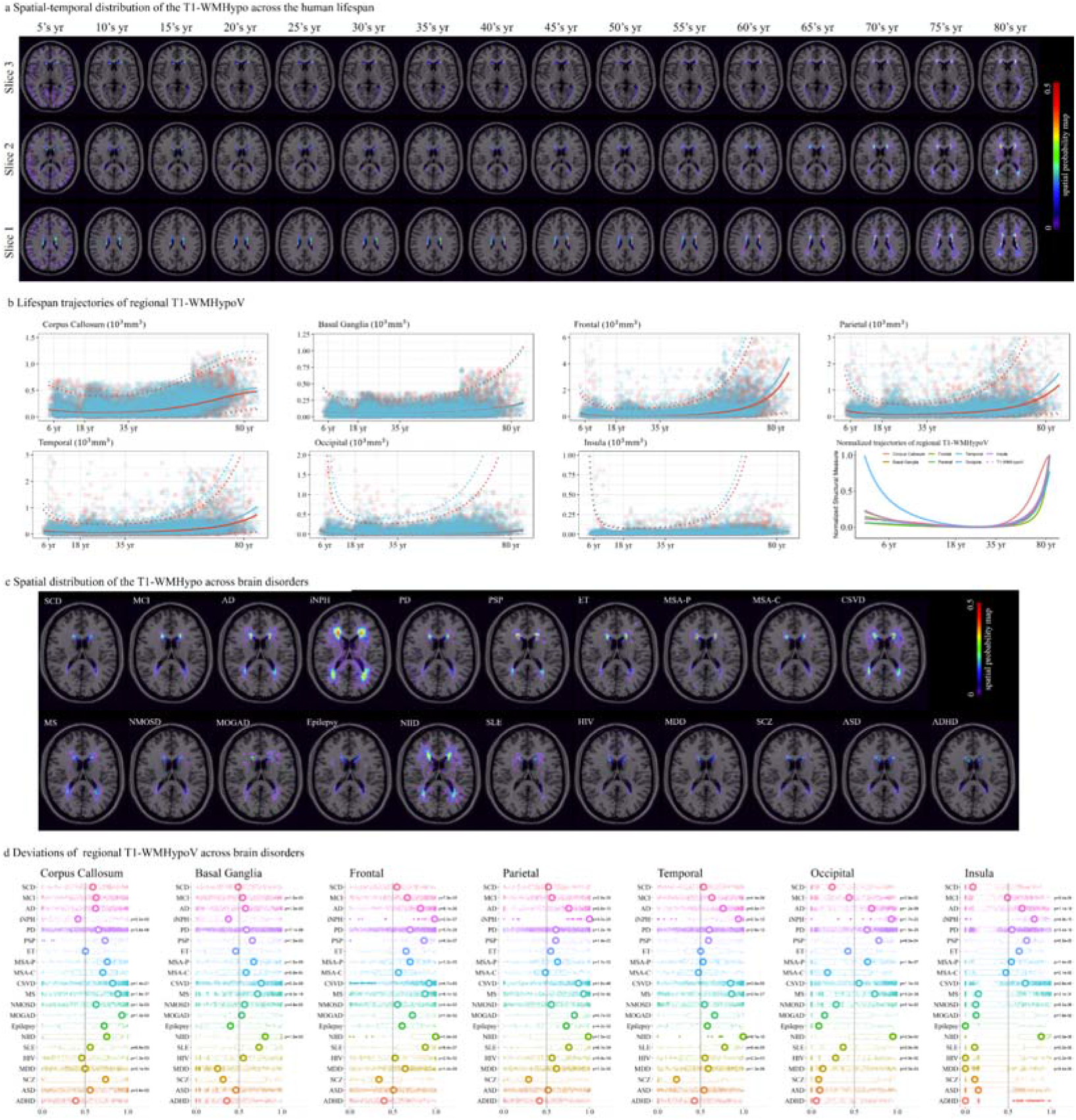
Spatial-temporal analyses of the T1-WMHypoV across HCs and brain disorders. (**a**) Spatial-temporal distribution of the T1-WMHypo, the spatial probability map was obtained by summing all the T1-WMHypo segmentation and divided by the case number within the age ranges; (**b**) Lifespan trajectories of regional T1-WMHypoV across seven brain regions; (**c**) Spatial distribution of T1-WMHypo across 21 brain disorders, the spatial probability map was obtained by summing all the T1-WMHypo segmentation and divided by the case number of the disorder; (**d**) Deviations of regional T1-WMHypoV across brain disorders and brain regions, the presented p values were FDR-corrected. Note: yr, years; T1-WMHypoV, white matter hypointensity on T1-weighted images; HC, healthy control; SCD, subjective cognitive decline; MCI, mild cognitive impairment; AD, Alzheimer’s disease; iNPH, idiopathic normal pressure hydrocephalus; PD, Parkinson’s disease; PSP, progressive supranuclear palsy; ET, essential tremor; MSA-P, multiple system atrophy - Parkinsonian variant; MSA-C, MSA-Cerebellar variant; CSVD, cerebral small vessel disease; MS, multiple sclerosis; NMOSD, neuromyelitis optica spectrum disorders; MOGAD, myelin oligodendrocyte glycoprotein antibody-associated disease; NIID, neuronal intranuclear inclusion disease; SLE, systemic lupus erythematosus; HIV, human immunodeficiency virus; MDD, major depressive disorder; SCZ, schizophrenia; ASD, autism spectrum disorder; ADHD, attention-deficit/hyperactivity disorder; FDR, false discovery rate.

### Additional Analyses to Enhance the Potential Value of White Matter Hypointensity

First, a subset of participants from HC, MCI, AD, CSVD, MS, and NMOSD had available longitudinal MRI scans (in this study, follow-up times for HCs and disease cases with MR scans ranged from 0.5 to 3 years, follow-up time points ranged from 1 to 4; HCs, n = 90, follow-up= 5.83-26.83 months; MCI, n = 47, follow-up= 5.83-25.73 months; AD, n = 98, follow-up= 6.00-25.57 months; CSVD, n = 95, follow-up= 10.57-37.00 months; MS, n = 72, follow-up= 5.80-30.73 months; and NMOSD, n = 68, follow-up= 5.47-29.27 months). Follow-up T1-WMHypoV values were first mapped together with baseline measurements across diagnostic groups (**Extended Data Fig. 4**). Longitudinal changes in T1-WMHypoV were then assessed using linear mixed-effect models, with follow-up time and diagnosis specified as fixed effects and participant as a random effect. The statistical significance of disease-specific longitudinal effects was corrected using FDR across diagnostic groups. Significant longitudinal increases in T1-WMHypoV were observed in MS (β = 0.32, 95% CI [0.09, 0.55], pFDR = 0.027) and NMOSD (β = 0.31, 95% CI [0.08, 0.54], pFDR = 0.027) over the disease course (**Extended Data Fig. 4)**. In contrast, no significant longitudinal changes were detected in MCI (β = 0.18, 95% CI [-0.13, 0.49], pFDR = 0.26), AD (β = 0.20, 95% CI [-0.06, 0.45], pFDR = 0.15), or CSVD (β = 0.20, 95% CI [0.00, 0.39], pFDR = 0.085), although the direction of effect in CSVD was positive.

Then, we repeated the analyses of T1-WMHypoV burden across HCs and brain disorders by raw T1-WMHypoV as well as correlation analyses with other raw brain structural metrics and clinical scores (**Extended Data Fig. 5**). The findings showed that the deviation score was more sensitive to reveal the T1-WMHypoV deviation in some brain disorders (e.g., PSP, MSA-P, Epilepsy and MDD). The deviation scores and raw values of T1-WMHypoV had comparable correlations with other brain tissue metrics and clinical scores, which further confirmed the findings by normative reference derived deviations.

## Discussion

This study established age- and sex-specific normative references for T1-WMHypoV across the human lifespan using a large international multicenter dataset and revealed its potential in both neurological and psychiatric disorders. (1) The derived lifespan trajectory follows a non-linear J-shaped pattern, with a modest decline from childhood to young adulthood and a progressive age-related increase beginning in midlife. (2) Using these normative references, patients across various disorders showed significantly greater T1-WMHypo burden than age- and sex-matched HCs. T1-WMHypo deviations could contribute to differential diagnosis, and elevated T1-WMHypo burden was consistently associated with brain atrophy, cognitive impairment, treatment response in PD, and disease progression in MS and NMOSD.

Together, these findings support T1-WMHypo deviations as a novel individualized imaging marker of WM pathophysiology independent of well-studied T2-FLAIR hyperintensity during development, aging and diseases, which should be highlighted in the clinic.

From a biological perspective, T1-WMHypo reflects heterogeneous pathophysiology substrates, including immature WM fibers, enlarged perivascular spaces, chronic demyelination, axonal loss, lacunar infarcts, and microangiopathy/arteriolosclerosis(*7–9*). Given this biological heterogeneity, interpreting the absolute burden of T1-WMHypo can be challenging, whereas normative modeling offers a principled approach for quantifying individual T1-WMHypo burden relative to population distributions. Using this framework, T1-WMHypoV exhibited a robust J-shaped trajectory across the lifespan, which was consistently observed across different WM regions, indicating a shared developmental and aging signature. The increasing limb of this trajectory in adulthood parallels previously reported normative patterns of T2-FLAIR defined WMHyper in adult populations from both Western and Chinese cohorts(*20, 21*). While these prior studies focused primarily on adulthood and reported progressive age-related increases in T2-WMHyper burden, our findings extend this perspective by characterizing T1-WMHypo across a much broader age range spanning early childhood to late adulthood, suggesting that both imaging markers may reflect partially overlapping biological processes related to cumulative WM injury during aging. Importantly, our results further revealed a decline in T1-WMHypo burden during childhood and adolescence, a phase rarely examined in previous studies of T1-WMHypo(*30*). This early-life decrease aligns with pediatric neuroimaging literature describing dynamic WM signal changes driven by ongoing myelination and tissue maturation, as well as enlarged perivascular space, indicating that similar T1WI signal alterations may arise from distinct biological substrates at different life stages(*18, 31–34*). T1-WMHypoV showed an inverse lifespan trajectory relative to total WMV and a concordant pattern with CSF volume in later life, linking increased T1-WMHypo burden to WM loss and ventricular expansion during brain aging(*23*).

Applying the normative framework across a wide range of neurological and psychiatric disorders revealed substantial heterogeneity in T1-WMHypo deviations. Across different brain disorders, the magnitude of deviation followed a severity continuum, with the largest effects observed in iNPH and NIID. These disorders are characterized by diffuse periventricular involvement, cerebrospinal fluid dysregulation, and progressive tissue rarefaction, consistent with extensive WM damage detectable on T1WIs(*35, 36*). High T1-WMHypo burden was also observed in conditions associated with diffuse WM injury and cerebral vascular pathology (e.g., enlarged perivascular space), including MS and CSVD. In MS, pronounced T1-WMHypo deviations also align with severe tissue injury beyond classical persistent “T1 black holes”, including chronic demyelination, axonal loss, matrix destruction, and gliosis, pathological features that contribute to reduced T1 signal intensity and reflect destructive lesion evolution and chronic WM damage (*37–39*). In contrast, NMOSD and MOGAD did not exhibit significant whole-brain T1-WMHypo deviations despite prominent lesion burden on T2-FLAIR imaging. This dissociation suggests that T1-WMHypo in such diseases may preferentially reflects more severe axonal damage and may aid in distinguishing inflammatory demyelinating disorders with different degrees of WM damage(*40, 41*). Moderate but consistent increases in T1-WMHypo burden were observed in neurodegenerative and systemic autoimmune conditions, including AD(*42*), PSP(*43*), and SLE(*44*), likely reflecting secondary WM degeneration accompanying cortical degeneration or systemic inflammatory processes. Modest yet statistically significant deviations were detected in MCI(*45*), PD(*46*), MSA-P(*47*), epilepsy(*48*), and MDD(*49*), i.e., in conditions where WM pathology is generally not considered the primary disease substrate. Collectively, these findings demonstrate that T1-WMHypo captures a shared yet selectively expressed dimension of underlying WM pathology across brain disorders.

Beyond disease-specific effects, T1-WMHypo showed consistent correlations with global brain structural metrics across both healthy individuals and patient groups, indicating close integration with large-scale brain integrity. Across various neurological disorders, higher T1-WMHypo burden was associated with reduced GMV, cortical thinning, subcortical and infratentorial involvement, and cerebrospinal fluid expansion, a pattern characteristic of widespread neurodegenerative processes rather than isolated WM pathology(*50–53*). In healthy individuals, these structural associations exhibited a clear age-dependent shift, with T1-WMHypo relating predominantly to WMV in younger adults and showing stronger coupling with CSF expansion and cortical thinning in later life. This transition suggests that T1-WMHypo evolves from a tissue-related feature in early adulthood to an atrophy-associated marker during aging, paralleling trajectories observed in neurodegenerative conditions(*54*). Importantly, many associations persisted after adjustment for T2-WMHyper burden, indicating that T1-WMHypo captures structural alterations not fully accounted for by conventional T2-FLAIR lesion load. Together, these correlation patterns support the interpretation that T1-WMHypo reflects a convergent process linking WM pathophysiology with GM degeneration and may provide complementary information for characterizing brain aging and disease-related atrophy.

T1-WMHypo had an additional contribution to a majority of disease differential diagnoses beyond multiple brain structural metrics and demographics, highlighting the benefit of including T1-WMHypo in clinical decision making, particularly if T2-FLAIR weighted imaging is unavailable. Meanwhile, it had consistent associations with clinical functional measures across various neurological disorders, particularly measures of global cognition and information processing speed, in disorders along the cognitive impairment spectrum, including MCI(*45*) and AD(*55*). Similar cognitive associations were also observed in other neurological conditions with recognized cognitive involvement, such as CSVD(*56*), MSA-C(*57*), and iNPH (*58*). Notably, comparable relationships were also detected in epilepsy(*59*), despite its distinct clinical phenotypes, suggesting that T1-WMHypo may capture a shared imaging correlate of cognitive dysfunction across diverse neurological conditions. Importantly, the association between cognitive impairment and T1-WMHypo remains after adjustment for T2-WMHyper burden, indicating that T1-WMHypo captures functional impairment independent of conventional T2-FLAIR lesion burden. In neuroinflammatory disorders, these associations were predominantly involved in processing speed-related measures, consistent with the established sensitivity of information processing efficiency to widespread WM disruption in demyelinating diseases(*60*). In contrast, motor- or mood-related measures showed limited associations with T1-WMHypoV across disorders, indicating relative domain specificity for cognitive function. Although previous studies have reported links between T1-WMHypo and age-related cognitive decline(*61*), the use of normative-based centile scores may attenuate cross-sectional brain-behavior correlations by accounting for age-related variance, thereby reducing associations driven by normal aging.

Our findings further indicate that T1-WMHypo burden carries prognostic relevance in various neurological disorders. Stratification based on T1-WMHypoV deviations showed that individuals with higher hypointensity burden experienced less favorable clinical outcomes, supporting its potential value for disease-specific risk stratification. In PD, patients with elevated T1-WMHypo burden exhibited more severe motor impairment and derived less benefit from pharmacological treatment, while showing limited additional improvement following DBS. These findings suggest that T1-WMHypo may constrain the functional reserve of motor networks, thereby modulating therapeutic responsiveness beyond primary dopaminergic deficits(*62*). In demyelinating disorders, T1-WMHypo burden was associated with disease progression. In MS, higher T1-WMHypo burden was linked to accelerated disability worsening and an increased risk of transition to secondary progressive disease, even after accounting for demographic factors and other brain structural metrics. This observation is consistent with the interpretation that T1-WMHypo reflects cumulative severe tissue disruption that may contribute to long-term neurological decline(*63, 64*). Additional longitudinal MRI analyses further showed progressive increases in T1-WMHypoV in MS and NMOSD, underscoring the disease-specific relevance of this marker.

To clarify the relationships between T1-WMHypoV and T2-WMHyperV, we investigated the correlation between T1-WMHypoV and T2-WMHyperV: they showed consistent patterns in aging but may result from different physiological changes during brain development and aging, and play distinct roles across the lifespan(*65*). The complementary clinical role of T1-WMHypo could be further supported by the clinical association in cases with low T2-WMHyper burdens, implying a partially independent role of T1-WMHypo in addition to T2-WMHyper in disease pathology. Spatial-temporal findings further displayed the spatial characteristic along the lifespan trajectory and brain disorders, supporting the main findings of whole brain T1-WMHypoV. They also revealed disease-specific T1-WMHypo spatial distribution, which indicated the disease-specific spatial involvement in brain disorders(*22*), while their underlying biological mechanisms should be further investigated.

Several limitations should be acknowledged. First, although the normative reference framework was derived from a large multicenter cohort, most analyses were cross-sectional. This design limits causal inference and constrains precise estimation of within-subject rates of T1-WMHypo progression. Future studies with denser longitudinal sampling will be essential to characterize individual trajectories and to validate the temporal sensitivity of T1-WMHypoV as a marker of disease evolution. Second, T1-WMHypoV was quantified using intensity-based segmentation of three-dimensional T1WIs. While this approach enables scalable and robust analysis across large datasets, T1-WMHypo reflects a heterogeneous mixture of pathological substrates. The absence of direct histopathological validation and limited multimodal integration preclude precise attribution of underlying biological mechanisms.

Integrating multimodal MRI and pathological correlation will be important for disentangling these components. Third, the present analyses primarily focused on global T1-WMHypo burden. Although additional regional and spatial analyses were performed, finer-grained compartmental modeling of WM may yield additional disease-specific insights and improve interpretability. Finally, the prognostic associations observed in only limited brain disorders, while encouraging, require external validation and prospective confirmation before clinical translation.

In summary, this study established age- and sex-specific normative references for T1-WMHypoV across the lifespan and demonstrated their applicability across a wide range of neurological and psychiatric disorders. By contextualizing individual T1-WMHypo burden within a normative framework, we show that this marker captures a dimension of WM pathophysiological changes partially independent of conventional T2-FLAIR hyperintensity. Normative deviation patterns vary substantially across diseases, contribute to disease diagnosis and relate to brain structural alterations, functional impairment, and clinical outcomes in various brain disorders. These findings highlight T1-WMHypoV deviations as an individualized imaging marker for characterizing WM pathophysiological characteristics and advancing precision approaches to brain development, aging and diseases.

## Methods

### Participants

Three-dimensional T1WI scans with isotropic voxel sizes of 1 mm or less were collected for a total of 41,735 scans. The dataset comprised 34,148 scans from HCs and 7,587 scans from individuals with neurological or psychiatric disorders. Detailed inclusion and exclusion criteria are provided as follows.

Inclusion criteria included (1) availability of high-quality three-dimensional T1-weighted MRI data, and (2) diagnoses established according to internationally recognized clinical standards. HCs were defined as individuals aged 0-100 years without any clinically diagnosed central nervous system disorders. Patient diagnoses were determined according to clinically established criteria.

Exclusion criteria included a history of additional neurological or neuropsychiatric disorders, incomplete MRI acquisition, inadequate image quality, or unsuccessful image segmentation. Follow-up scans were excluded from cross-sectional analyses. After quality control and exclusions, the final cohort comprised 34,148 HCs and 7,587 patients (details see **Table 1**). The distribution of participants across 146 centers is shown in **Fig. 1a,b**.

Age or date of birth was extracted from DICOM metadata or obtained directly from participants or their legal guardians. Available clinical variables included age, sex, and years of education (n=6,442). Cognitive assessments included MoCA (n=5,636) and MMSE (n=4,850). Additional cognitive measures were available in subsets of MS, NMOSD and MOGAD patients, including SDMT (n=950) and PASAT (n=926). Motor and disability assessments were available for specific patient groups. Among PD patients, UPDRS-III assessments were obtained in the off-medication state (n=788), on-medication state (n=783), and after DBS (n=498). Baseline and follow-up EDSS scores were available for 850 MS and 394 NMOSD patients, respectively, for neuroinflammatory diseases. Mood assessment, including Hamilton Anxiety Rating Scale (HAMA, n=1,866) and Hamilton Depression Rating Scale (HAMD, n=1,873) were assessed for specific patient groups. Pittsburgh Sleep Quality Index (POSI, n=253) for a subset was also acquired.

### MRI Acquisition and Image Processing

MRI data were obtained from 146 sites. All participants underwent three-dimensional T1WIs, typically using a magnetization-prepared rapid gradient echo sequence, with isotropic voxel sizes of 1 mm or less. A subset of participants additionally underwent T2-weighted FLAIR imaging. All images were independently reviewed for quality by two trained raters, with discrepancies resolved by consensus involving a third reviewer. T1WI data were processed using the FreeSurfer (version 7.3.2). Standard preprocessing steps included correction for intensity non-uniformity, skull stripping, intensity normalization, and registration to Talairach space. Volumetric measures were extracted from the aseg.stats and aparc.stats outputs, including WMV, cortical GMV, sGMV, cerebellar volume, brainstem volume, CSF volume, mean cortical thickness, total cortical surface area, and TIV. T1-WMHypoV was derived from intensity-based segmentation within the WM mask as implemented in FreeSurfer. All T1-WMHypo segmented images were spatially resampled and aligned to a standard template before regional analysis. WMH were parcellated into seven anatomical regions using the Hammers atlas: corpus callosum, basal ganglia, frontal, parietal, temporal, occipital and insula. For participants with available T2-weighted FLAIR images, T2-WMHyper segmentation was performed using a previously validated deep learning model(*66*).

Quality control was conducted at multiple stages of the processing pipeline. Raw MRI scans were visually inspected for motion artifacts and acquisition-related abnormalities. Automated outlier detection was applied to segmentation outputs, and manual inspection was performed when necessary. To mitigate residual site-related variability, values exceeding three standard deviations from the site-specific mean were excluded from subsequent analyses.

### Normative modeling

Normative lifespan models for T1-WMHypoV and other brain structure metrics were constructed using the HC cohort within the GAMLSS implemented in R (version 4.3). Age and sex were included as fixed effects, and scanning site/center was modeled as a random effect. A generalized gamma distribution was selected to account for the right-skewed distribution of volume measures. Age-related effects were modeled using fractional polynomials, with model complexity determined by the Bayesian Information Criterion. To reduce potential bias in deviation estimation, 10-fold cross-validation was applied. Individualized centile scores were derived for each participant based on the normative models. All patient scans were acquired at sites represented in the normative dataset, allowing appropriate adjustment for site-related effects. Age-related rates of change were estimated by computing the first derivative of the fitted median trajectory across the lifespan.

### Statistical analysis

All statistical analyses were performed using R software (version 4.3, R Foundation for Statistical Computing, Vienna, Austria). Categorical variables are presented as counts (percentages) and were compared using Pearson’s chi-square tests. Continuous variables are reported as means ± standard deviation and compared using one-way analysis of variance when approximately normally distributed, or as medians with interquartile ranges (or ranges) and compared using the Kruskal-Wallis test when distributions were non-normal.

Normative deviation scores of T1-WMHypoV and other structure metrics were calculated for all participants based on the age- and sex-specific normative reference models. Group-level differences in deviations T1-WMHypoV and other brain structure metrics between patients and HCs were assessed using Monte Carlo permutation tests with the Welch t statistic. When available, HCs were matched to patients by age, sex, and scanning site; otherwise, unmatched HCs were used. Effect sizes were quantified using Cohen’s d with 95%CIs. Multiple comparisons across disease groups were controlled using the FDR procedure.

To assess whether the T1-WMHypoV has an additional contribution to the disease differential diagnosis, we performed a series of classifications among the above brain disorders using logistic regression. To test the potential contribution of T1-WMHypoV deviations, we compared the adjusted McFadden R² between full (all variates were considered) and reduced models (all variates except T1-WMHypoV were considered) for each pair of disorder groups. Likelihood Ratio Test was adopted to test the model performances between each paired full and reduced models, a p < 0.05 was considered statistically significant. McFadden R²of both full and reduced models, the absolute (defined as [McFadden R² of full model - McFadden R² of reduced model]) and relative (defined as [McFadden R² of full model - McFadden R² of reduced model]/McFadden R² of reduced model*100%) changes of McFadden R², and the p values distribution of Likelihood Ratio Test across the paired classifications among brain disorders were reported.

Associations between T1-WMHypoV deviations with brain structural metrics and clinical scores were examined using Pearson correlation analyses. Three complementary approaches were applied: univariate correlations, partial correlations adjusted for age, sex, and TIV, and partial correlations with additional adjustment for T2-WMHyperV to assess independence from conventional lesion burden. Brain structural metrics included T2-WMHyperV, WMV, cortical and sGMVs, cerebellar and brainstem volumes, CSF volume, mean cortical thickness, and total cortical surface area. Clinical scores comprised global cognitive scores, disability and motor assessments, and affective and sleep-related scales. Multiple comparisons were controlled using FDR correction across diagnostic groups and outcome measures. To evaluate the prognostic value of T1-WMHypoV deviations, analyses were conducted in PD, MS and NMOSD. For treatment response assessment in PD, we defined the PD cases with extreme deviations (centile score > 0.95) as the high T1-WMHypo burden subgroup, and the remaining as the low T1-WMHypo burden subgroup. Between-group differences on motor improvement after medication and one-year follow-up after DBS therapy were analyzed with Student’s t-test. Above deviation cutoffs were also applied for MS and NMOSD cases to define the high and low T1-WMHypo burden subgroups. Kaplan-Meier analysis with a log-rank test was conducted to investigate the potential differences in disease progression of the stratified subgroups. We additionally conducted multivariate Cox proportional hazards regression using the deviation scores to test the independent predictive value of T1-WMHypoV deviations, by considering age, sex, and other brain structural metrics.

Based on the available T2-WMHyperV, we analyzed the correlations between absolute raw T1-WMHyoV and T2-WMHyperV by univariate Pearson correlation across HC and brain disorder groups. Then we fitted the normative reference of T2-WMHyperV with the same method as described in the T1-WMHypoV normative reference construction. The deviations of T2-WMHyperV across HCs and brain disorders were calculated and compared as described in the T1-WMHypoV deviation analysis. Univariate Pearson correlation with other brain structural metrics and clinical scores was conducted to test the clinical relevance of T2-WMHyperV deviations for comparison with those in T1-WMHypoV.

Longitudinal analyses were conducted in a subset of HCs and patients with available follow-up MRI scans. Linear mixed-effects models were applied, with follow-up time, diagnosis, and their interaction specified as fixed effects and participant included as a random effect. Statistical significance of diagnosis-specific longitudinal effects was corrected for multiple comparisons using FDR across disease groups.

All statistical tests were two-sided, and statistical significance was defined as P < 0.05.

### Human Ethics and Consent to Participate declarations

This study was conducted in accordance with the Declaration of Helsinki and complied with the Strengthening the Reporting of Observational Studies in Epidemiology (STROBE) guidelines. Ethical approval was obtained from the Ethics Committees of Beijing Tiantan Hospital, Capital Medical University (approval numbers: KY-2019-050-02, KY-2019-140-02, KY-2024-221-02, and KY-2025-203-03; Clinical Trial Number: NCT06643858). Written informed consent was obtained from all participants or their legally authorized representatives.

## Data availability

A part of brain morphological imaging phenotype measures (example datasets approved by local centers) derived by FreeSurfer have been shared at GitHub (https://github.com/zhuozhizheng/T1-WMHypo-normative-references) and Science Data Bank (https://www.scidb.cn/s/yqy6Jb).

## Code availability

Main codes used for the statistical analyses and figures are publicly available at GitHub (https://github.com/zhuozhizheng/T1-WMHypo-normative-references) and Science Data Bank (https://www.scidb.cn/s/yqy6Jb).

## Acknowledgments

We acknowledge all the contributors of Chinese Brain Health Imaging Consortium (CHART) who provided MR data from healthy volunteers.

This work was supported by a grant from the National Natural Science Foundation of China (NO. 82572171) (ZZ), National Key R&D Program of China (NO. 2025YFC3609700) (YL), the Chinese Institutes for Medical Research, Beijing (NO. CX23YZ01) (YL), the STI 2030-the major projects of the Brain Science and Brain-Inspired Intelligence Technology (2021ZD0200500) (XNZ), the Key Program of the National Natural Science Foundation of China (NO. 82330057) (YL), The Cross-Research Project of Beijing Science and Technology Star Program (NO. 20230484428) (YL), the Beijing Outstanding Young Scientist Program (JWZQ20240101025) (YL), Beijing Municipal Natural Science Foundation for Distinguished Young Scholars (No. JQ20035) (YL), Beijing Hospital Management Center-Climb Plan (No. DFL20220503) (YL), Beijing Young Scholars (YL), the Major Fund for International Collaboration of National Natural Science Foundation of China (81220108014) (XNZ) and the National Basic Science Data Center ‘Interdisciplinary Brain Database for In vivo Population Imaging’ (ID-BRAIN) (XNZ).

## Author contributions

Study design: Zhizheng Zhuo, Shan Lv, Yaou Liu.

Data analysis: Zhizheng Zhuo, Renlong Zhang, Shan Lv, Xiaozhou Du.

Statistical analysis: Zhizheng Zhuo, Shan Lv, Xiaozhou Du.

Data collection: Zhizheng Zhuo, Shan Lv, Peng Gao, Ningnannan Zhang, Li Chai, Xiaozhou Du, Yuna Li, Yinshan Wang, Ye He, Ping Wang, Zixuan Lin, Shuiwei Xia, Yunyun Duan, Wei Wang, Yunling Wang, Jiansong Ji, and Chinese Brain Health Imaging Consortium (CHART).

Manuscript writing: Zhizheng Zhuo, Shan Lv.

Manuscript editing: Shan Lv, Peng Gao, Li Chai, Ningnannan Zhang, Xiaozhou Du, Dan Cheng, Yuna Li, Wei Wang, Yunling Wang, Jiansong Ji, James Cole, Dennis M. Hedderich, Xi-Nian Zuo and Yaou Liu.

Other Contributors of Chinese Brain Health Imaging Consortium (CHART) who are not listed in the author list.

Zhanjun Zhang^1^, Yaojing Chen^1^, Panjie Yan^1^, Mengjie Rong^1^, Liping Ai^2^, Xinxiang Zhao^2^, Feng Ao^3^, Yalin Bai^4^, Zheqi Gao^4^, Yutong Bai^5^, Jianguo Zhang^5^, Haihua Bao^6^, Ya Guo^6^, Qing Cai^7^, Chu-Chung Huang^7^, Jiancang Cao^8^, Lianping Zhao^8^, Feiyan Chen^9^, Hongjian He^9^, Feng Chen^10^, Fei Wang^10^, Kun Chen^11^, Haiyan Wu^11^, Zaixu Cui^12^, Yang Li^12^, Hui Dai^13^, Demao Deng^14^, Yuhui Du^15^, Guoguang Fan^16^, Ying Fu^17,18^, Lubin Gou^19,20^, Junqiang Lei^19,20^, Chunjie Guo^21^, Xuemei Han^22^, Yang Hou^23^, Basheng Hu^24^, Yuankui Wu^24^, Jianping Hu^25^, Peiyu Huang^26^, Hui-Jie Li^27^, Jianrui Li^28^, Zhiqiang Zhang^28^, Junlin Li^29^, SongTao Yin^29^, Kexuan Li^30^, Zhenyu Shu^30^, Shaowu Li^31^, Lanxi Meng^31^, Wenling Li^32^, Li Zhu^32^, Xinyu Li^33^, Cheng Xu^33^, Yao Li^34^, Jie Luo^34^, Yongmei Li^35^, Yunfei Li^36^, Xiaohu Zhao^36^, Peipeng Liang^37^, Haiyan Liao^38^, Qin Shen^39^, Ching-Po Lin^39,40^, Jun Liu^41,42^, Jungang Liu^43^, Weiqi Liu^44^,

Ying Liu^45^, Jun Qiu^45^, Shanshan Lu^46^, Feiyun Wu^46^, Su Lui^47^, Hailin Ma^48^, Ning Mao^49^, Linghui Meng^50^, Jian Zhao^50^, Yinfeng Qian^51^, Jiajia Zhu^51^, Juwei Shao^52^, Decai Tian^53^, Yilong Wang^53^, Jun Xu^53^, Yiheng Tu^54^, Jinhui Wang^55,56,57^, Junkai Wang^58^, Wen Wang^59^,

Xiaochun Wang^60^, Feifei Zhang^60^, Yong Wang^61^, Zheng Wang^62,63^, Jinyuan Weng^64^, Ying Wu^65^, Xiaona Xia^66^, Xiangshui Meng^66^, Jiaheng Xu^67^, Tong Zhang^67^, Rui Xu^68^, Ruimeng Yang^69,70^, Xi Yin^71^, Yulong Yin^72^, Xin Zhao^72^, Rongjun Yu^73,74^, Zhen Yuan^75^, Hailian Zhang^76^, Yaqin Zhang^77^, Zhihua Zhang^78^, Xia Zhe^79^, Fuqing Zhou^80^, Kuncheng Li^81^, Xujun Duan^82^

1. State Key Laboratory of Cognitive Neuroscience and Learning, Faculty of Psychology, Beijing Normal University, Beijing, China.
2. Department of Radiology, The Second Affiliated Hospital of Kunming Medical University, Kunming, China.
3. Department of Radiology, Renmin Hospital, Hubei University of Medicine, Shiyan, China.
4. Department of Radiology, Yuci Renmin Hospital, Jinzhong, China.
5. Department of Neurosurgery, Beijing Tiantan Hospital, Capital Medical University, Beijing, China.
6. Department of Medical Imaging Center, Qinghai University Affiliated Hospital, Xining, China.
7. School of Psychology and Cognitive Science, East China Normal University, Shanghai, China.
8. Department of Radiology, Gansu Provincial Hospital, Lanzhou, China.
9. School of Physics, Zhejiang University, Hangzhou, China.
10. Department of Radiology, Hainan General Hospital, Haikou, China.
11. Centre for Cognitive and Brain Sciences and Department of Psychology, University of Macau, Macau, China.
12. Chinese Institute for Brain Research, Beijing, China.
13. Department of Radiology, Affiliated Hospital of Zunyi Medical University, Zunyi, China.
14. Department of Radiology, The People’s Hospital of Guangxi Zhuang Autonomous Region, Nanning, China.
15. School of Computer and Information Technology, Shanxi University, Taiyuan, China.
16. Department of Radiology, The First Hospital of China Medical University, Shenyang, China.
17. Department of Neurology and Institute of Neurology, The First Affiliated Hospital of Fujian Medical University, Fuzhou, China.
18. Institute of Neuroscience and Fujian Key Laboratory of Molecular Neurology, Fujian Medican University, Fuzhou, China.
19. Department of Radiology, Lanzhou University First Affiliated Hospital, Lanzhou, China.
20. Gansu Province Clinical Research Neurology, Center for Radiology Imaging, Lanzhou, China.
21. Department of Radiology, The First Hospital of Jilin University, Changchun, China.
22. Department of Neurology, China-Japan Union Hospital of Jilin University, Changchun, China. China– Japan Union Hospital of Jilin University, Changchun, China.
23. Department of Radiology, Shengjing Hospital of China Medical University, Shenyang, China.
24. Department of Radiology, Nanfang Hospital, Southern Medical University, Guangzhou, China.
25. Department of Radiology, The First Affiliated Hospital of Fujian Medical University, Fuzhou, China.
26. Department of Radiology, The Second Affiliated Hospital of Zhejiang University School of Medicine, Hangzhou, China.
27. State Key Laboratory of Cognitive Science and Mental Health, Institute of Psychology, Chinese Academy of Sciences, Beijing, China.
28. Department of Diagnostic Radiology, Affiliated Jinling Hospital, Medical School of Nanjing University, Nanjing, China.
29. Department of Imaging Medicine, Inner Mongolia Autonomous Region People’s Hospital, Hohhot, China.
30. Department of Radiology, Zhejiang Province People’s Hospital, Hangzhou, China.
31. Beijing Neurosurgical Institute, Beijing, China.
32. Department of Radiology, General Hospital of Ningxia Medical University, Yinchuan, China.
33. Department of Imaging, Shanxi Provincial People’s Hospital, Taiyuan, China.
34. National Engineering Research Center of Advanced Magnetic Resonance Technologies for Diagnosis and Therapy, School of Biomedical Engineering, Shanghai Jiao Tong University, Shanghai, China.
35. Department of Radiology, The First Affiliated Hospital of Chongqing Medical University, Chongqing, China.
36. Department of Radiology, The Fifth People’s Hospital of Shanghai Fudan University, Shanghai, China.
37. School of Psychology, Capital Normal University, Beijing, China.
38. Department of Radiology, The Second Xiangya Hospital, Central South University, Changsha, China.
39. Institute of Neuroscience, National Yang Ming Chiao Tung University, Taipei, Taiwan.
40. Department of Education and Research, Taipei City Hospital, Taipei, Taiwan.
41. Department of Radiology, Chongqing University Central Hospital, Chongqing, China.
42. Department of Radiology, Chongqing Emergency Medical Center, Chongqing, China.
43. Department of Radiology, Xiamen Children’s Hospital (Children’s Hospital of Fudan University at Xiamen), Xiamen, China.
44. Sophmind Technology (Beijing), Beijing, China.
45. Department of Radiology, The First Affiliated Hospital of USTC, University of Science and Technology of China, Hefei, China.
46. Department of Radiology, The First Affiliated Hospital with Nanjing Medical University, Nanjing, China.
47. Huaxi MR Research Center (HMRRC), Department of Radiology, West China Hospital of Sichuan University, Chengdu, China.
48. Tibet Autonomous Region Key Laboratory for High Altitude Brain Science and Environmental Acclimatization, Lhasa, China.
49. Department of Radiology, Yantai Yuhuangding Hospital, Qingdao University, Yantai, China.
50. Department of Radiology, Hebei Medical University Third Hospital, Shijiazhuang, China.
51. Department of Radiology, The First Affiliated Hospital of Anhui Medical University, Hefei, China.
52. The Affiliated Hospital of Yunnan University, Kunming, China.
53. Department of Neurology, Beijing Tiantan Hospital, Capital Medical University, Beijing, China.
54. Department of Psychology, University of Chinese Academy of Sciences, Beijing, China.
55. Institute for Brain Research and Rehabilitation, South China Normal University, Guangzhou, China.
56. Key Laboratory of Brain, Cognition and Education Sciences (Ministry of Education), South China Normal University, Guangzhou, China.
57. Center for Studies of Psychological Application, South China Normal University, Guangzhou, China.
58. Department of Radiology, Aerospace Center Hospital, Beijing, China.
59. Department of Radiology and Functional and Molecular Imaging Key Lab of Shaanxi Province, Fourth Military Medical University, Xi’an, China.
60. Department of Radiology, First Hospital of Shanxi Medical University, Taiyuan, China.
61. Department of Radiology, The First Hospital of Hebei Medical University, Shijiazhuang, China.
62. School of Psychological and Cognitive Sciences, Peking University, Beijing, China.
63. School of Biomedical Engineering, Hainan University, Sanya, China.
64. Philips (China) Investment, Shanghai, China.
65. School of Medical Imaging, Shanxi Medical University, Taiyuan, China.
66. Department of Radiology, Qilu Hospital (Qingdao), Cheeloo College of Medicine, Shandong University, Qingdao, China.
67. Department of Radiology, The Fourth Affiliated Hospital of Harbin Medical University, Harbin, China.
68. Department of Radiology, First Affiliated Hospital of Xinjiang Medical University, Urumqi, China.
69. Department of Radiology, The Second Affiliated Hospital, South China University of Technology, Guangzhou, China.
70. Department of Radiology, Guangzhou First People’s Hospital, Guangzhou, China.
71. Medical Imaging Center, The First Affiliated Hospital of Shihezi University, Shihezi, China.
72. School of Psychology, Northwest Normal University, Lanzhou, China.
73. Life Science Imaging Center, Hong Kong Baptist University, Hong Kong, China.
74. Department of Education and Psychology, Academy of Wellness and Human Development, Faculty of Arts and Social Sciences, Hong Kong Baptist University, Hong Kong, China.
75. Faculty of Health Sciences, University of Macau, Macau, China.
76. Department of Radiology, The Fifth People’s Hospital of Qinghai Province, Xining, China.
77. Department of Radiology, The Fifth Affiliated Hospital, Sun Yat-sen University, Zhuhai, China.
78. Department of Neurosurgery, Tsinghua University Yuquan Hospital (Tsinghua University Hospital of Integrated Traditional Chinese and Western Medicine), Beijing, China.
79. Department of MRI, Shaanxi Provincial People’s Hospital, Xi’an, China.
80. Department of Radiology, The First Affiliated Hospital, Nanchang University, Nanchang, China.
81. Department of Radiology, Xuanwu Hospital, Capital Medical University, Beijing, China.
82. The Clinical Hospital of Chengdu Brain Science Institute, School of Life Science and Technology, University of Electronic Science and Technology of China, Chengdu, China.

## Extended data Figures

**Extended data Fig.1.**
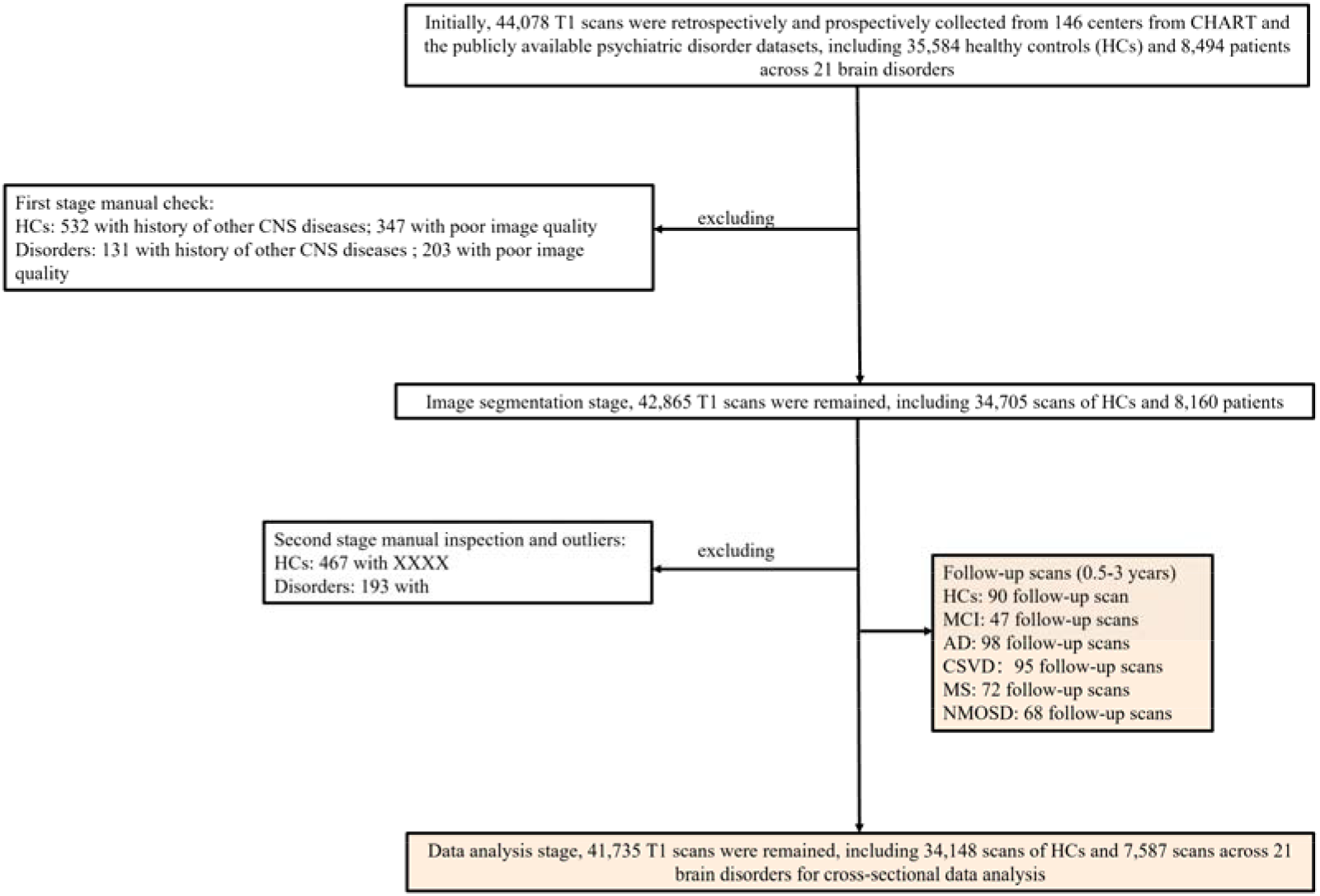
A brief description of the cases included in this study. Note: HC, healthy controls; MCI, mild cognitive impairment; AD, Alzheimer’s disease; CSVD, cerebral small vessel disease; MS, multiple sclerosis; NMOSD, neuromyelitis optica spectrum disorders.

**Extended data Fig.2.**
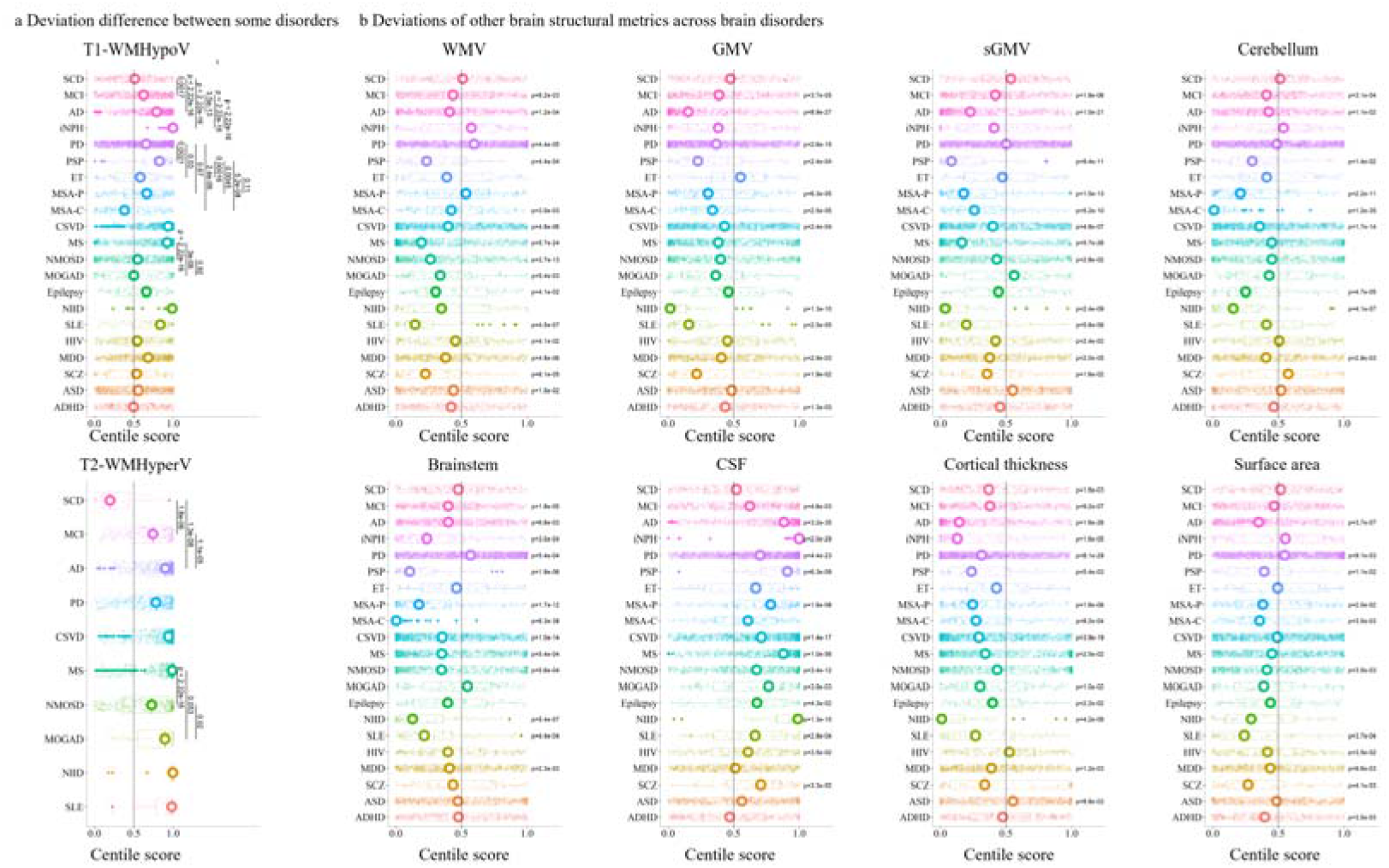
Between-group difference of T1-WMHypoV and T2-WMHyperV deviations, and deviations of other brain structural metrics. Note: T1-WMHypoV, white matter hypointensity on T1-weighted images; T2-WMHyperV, white matter hyperintensity on T2/FLAIR images; SCD, subjective cognitive decline; MCI, mild cognitive impairment; AD, Alzheimer’s disease; iNPH, idiopathic normal pressure hydrocephalus; PD, Parkinson’s disease; PSP, progressive supranuclear palsy; ET, essential tremor; MSA-P, multiple system atrophy - Parkinsonian variant; MSA-C, MSA-Cerebellar variant; CSVD, cerebral small vessel disease; MS, multiple sclerosis; NMOSD, neuromyelitis optica spectrum disorders; MOGAD, myelin oligodendrocyte glycoprotein antibody-associated disease; NIID, neuronal intranuclear inclusion disease; SLE, systemic lupus erythematosus; HIV, human immunodeficiency virus; MDD, major depressive disorder; SCZ, schizophrenia; ASD, autism spectrum disorder; ADHD, attention-deficit/hyperactivity disorder; GMV, gray matter volume; sGMV, subcortical gray matter volume; WMV, white matter volume; CSF, cerebrospinal fluid.

**Extended data Fig.3.**
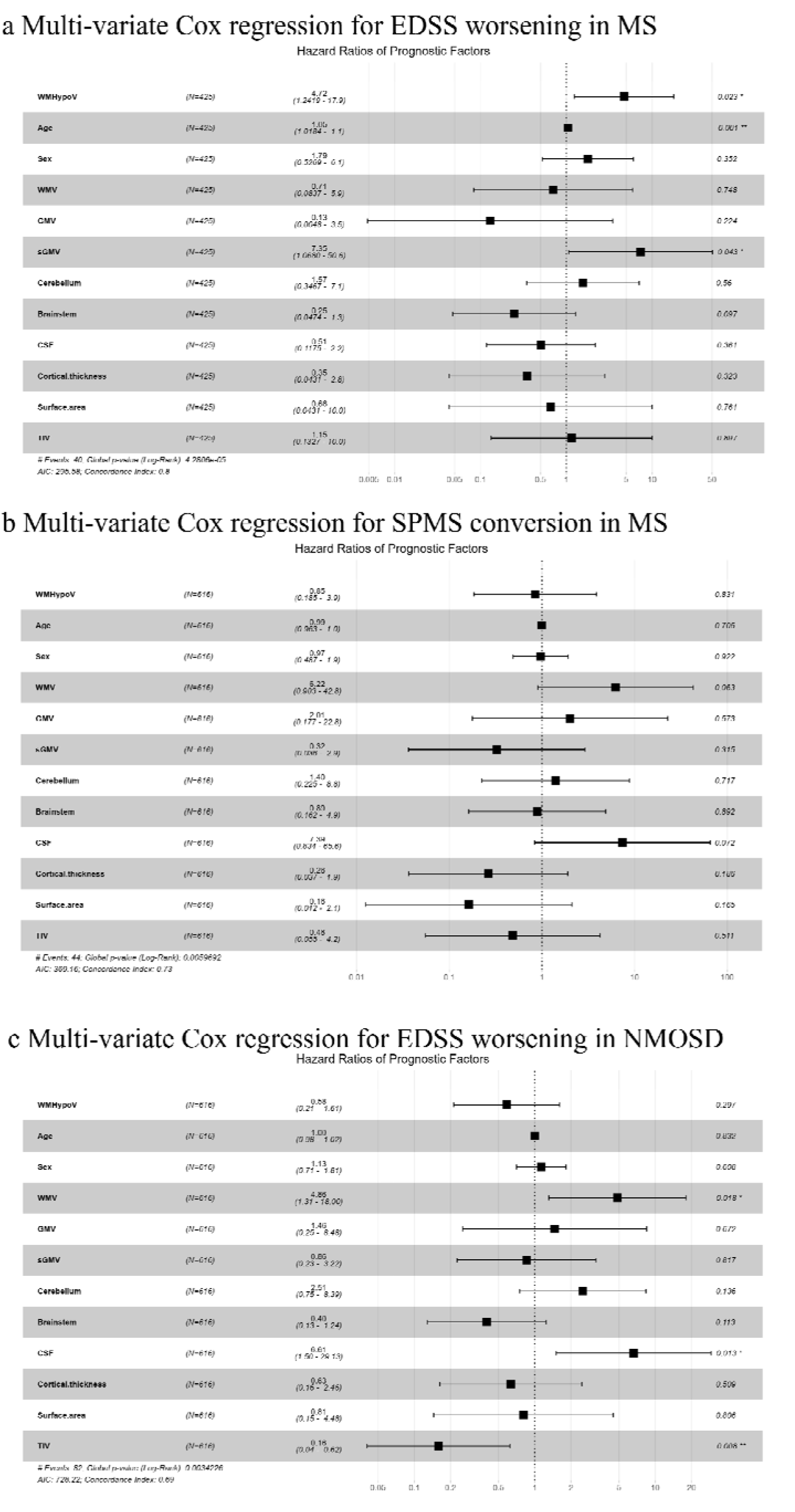
Multivariate Cox-regression for disease progression in neuroinflammatory diseases. Note: T1-WMHypoV, white matter hypointensity on T1-weighted images; T2-WMHyperV, white matter hyperintensity on T2-FLAIR images; MS, multiple sclerosis; NMOSD, neuromyelitis optica spectrum disorders; GMV, gray matter volume; sGMV, subcortical gray matter volume; WMV, white matter volume; CSF, cerebrospinal fluid; TIV, total intracranial volume; EDSS, Expanded Disability Status Scale; SPMS, Secondary Progressive MS.

**Extended data Fig.4.**
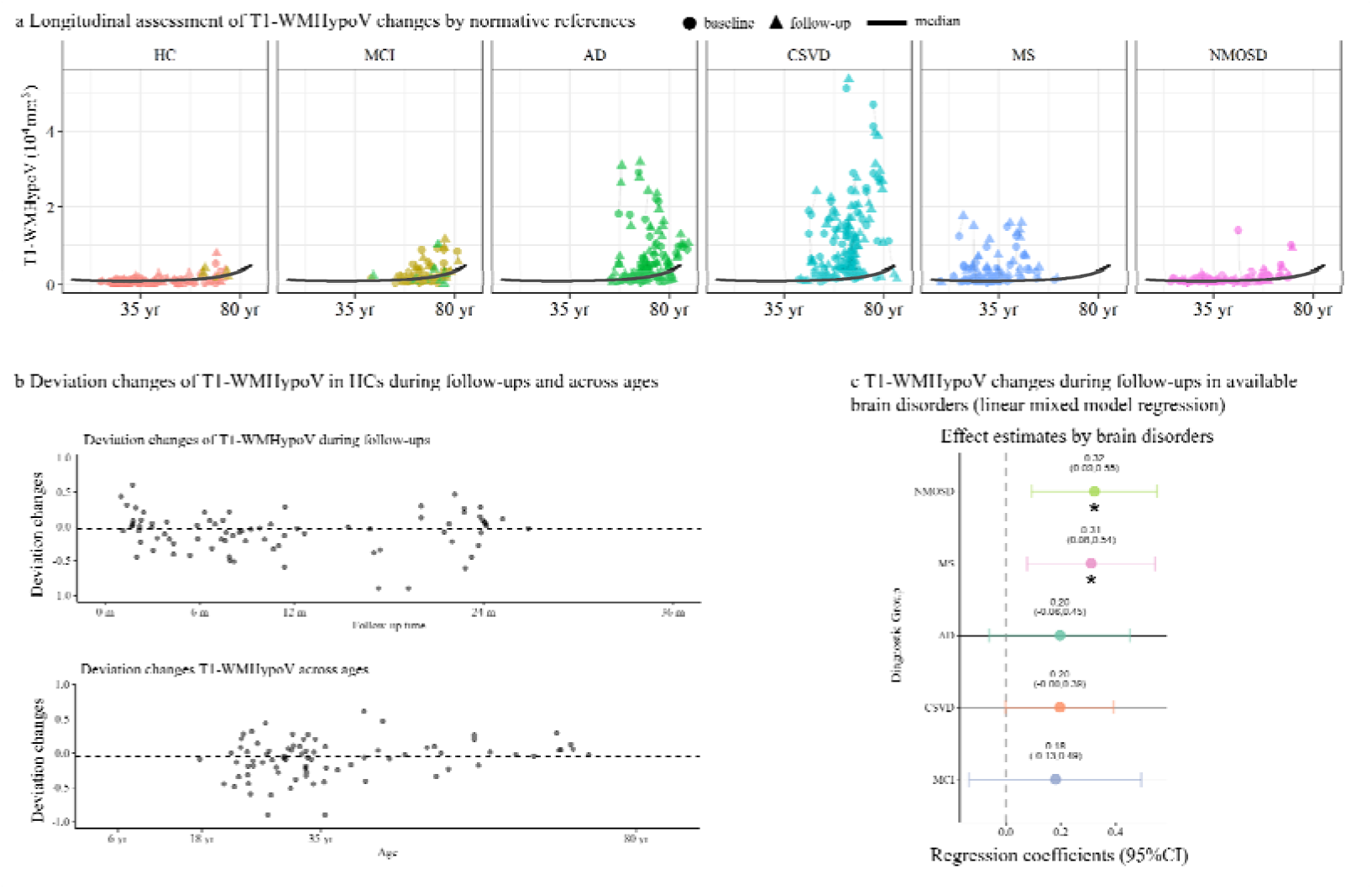
Longitudinal assessment of HCs and brain disorders. (**a**) Follow-up T1-WMHypoV of HCs and patients with brain disorders in the framework of normative references; (**b**) Follow-up T1-WMHypoV deviations of HCs across follow-up times and ages; (**c**) The T1-WMHypoV deviation score changes of follow-up scans of diseases in a linear mixed model. Note: T1-WMHypoV, white matter hypointensity on T1-weighted images; m, months; yr, years; HC, healthy control; MCI, mild cognitive impairment; AD, Alzheimer’s disease; CSVD, cerebral small vessel disease; MS, multiple sclerosis; NMOSD, neuromyelitis optica spectrum disorder.

**Extended data Fig.5.**
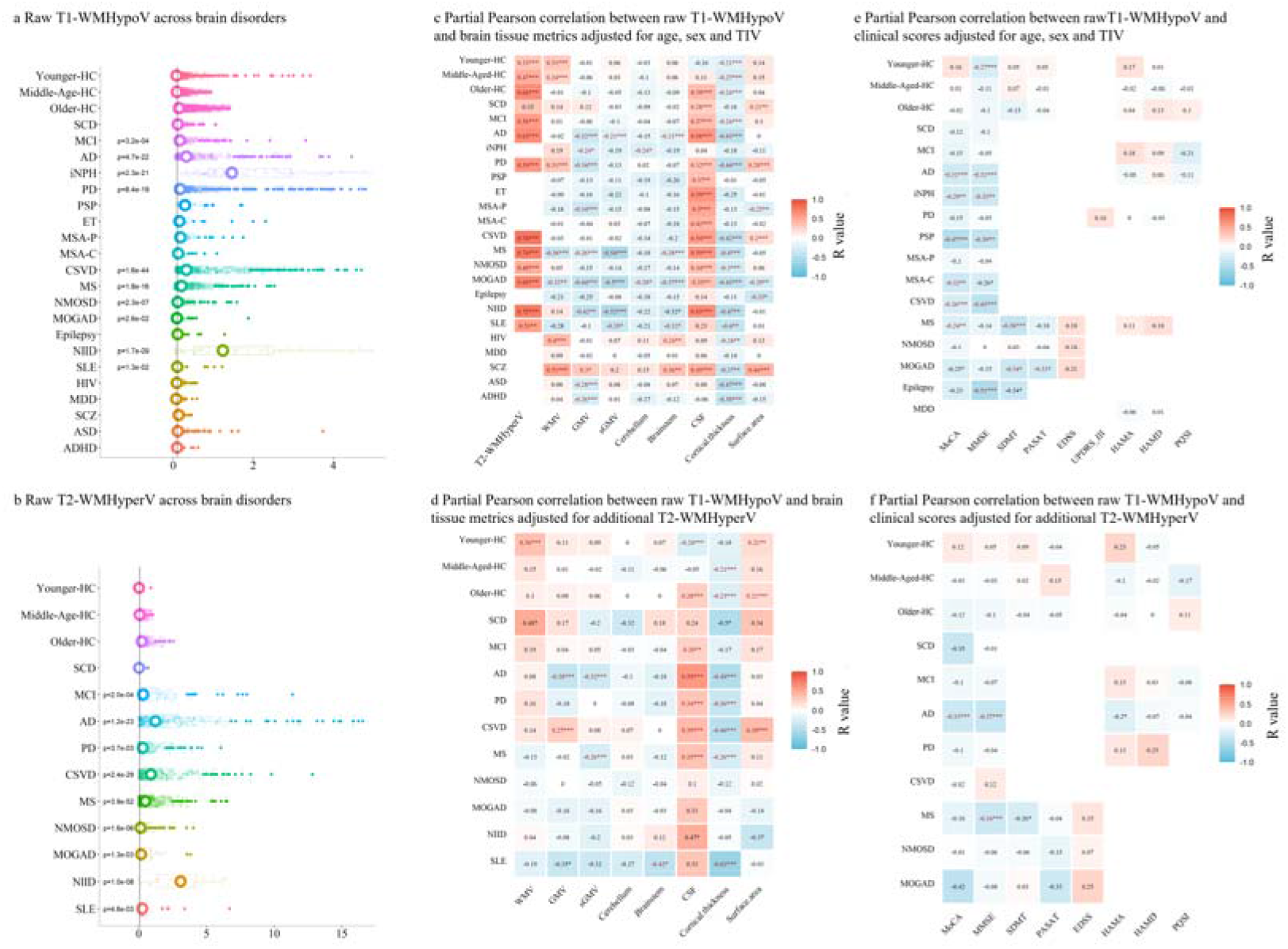
Findings by raw T1-WMHypoV and T2-WMHyperV. Note: T1-WMHypoV, white matter hypointensity on T1-weighted images; T2-WMHyperV, white matter hyperintensity on T2/FLAIR images; HC, healthy control; SCD, subjective cognitive decline; MCI, mild cognitive impairment; AD, Alzheimer’s disease; iNPH, idiopathic normal pressure hydrocephalus; PD, Parkinson’s disease; PSP, progressive supranuclear palsy; ET, essential tremor; MSA-P, multiple system atrophy - Parkinsonian variant; MSA-C, MSA-Cerebellar variant; CSVD, cerebral small vessel disease; MS, multiple sclerosis; NMOSD, neuromyelitis optica spectrum disorders; MOGAD, myelin oligodendrocyte glycoprotein antibody-associated disease; NIID, neuronal intranuclear inclusion disease; SLE, systemic lupus erythematosus; HIV, human immunodeficiency virus; MDD, major depressive disorder; SCZ, schizophrenia; ASD, autism spectrum disorder; ADHD, attention-deficit/hyperactivity disorder; GMV, gray matter volume; sGMV, subcortical gray matter volume; WMV, white matter volume; CSF, cerebrospinal fluid; TIV, total intracranial volume; T2-WMHyperV, white matter hyperintensity on T2/FLAIR images; MoCA, Montreal Cognitive Assessment; MMSE, Mini-Mental State Examination; SDMT, Symbol Digit Modalities Test; PASAT, Paced Auditory Serial Addition Test; EDSS, Expanded Disability Status Scale; UPDRS-III, Unified Parkinson’s Disease Rating Scale - Part III; HAMA, Hamilton Anxiety Rating Scale; HAMD, Hamilton Depression Rating Scale; PQSI, Pittsburgh Sleep Quality Index; DBS, deep brain stimulation. * p< 0.05, ** p< 0.005, *** p< 0.001

