## Extended data Fig.1 for "Unraveling the Lifespan Trajectory of White Matter Hypointensity on T1-weighted MR Images and Clinical Application in Various Brain Disorders"

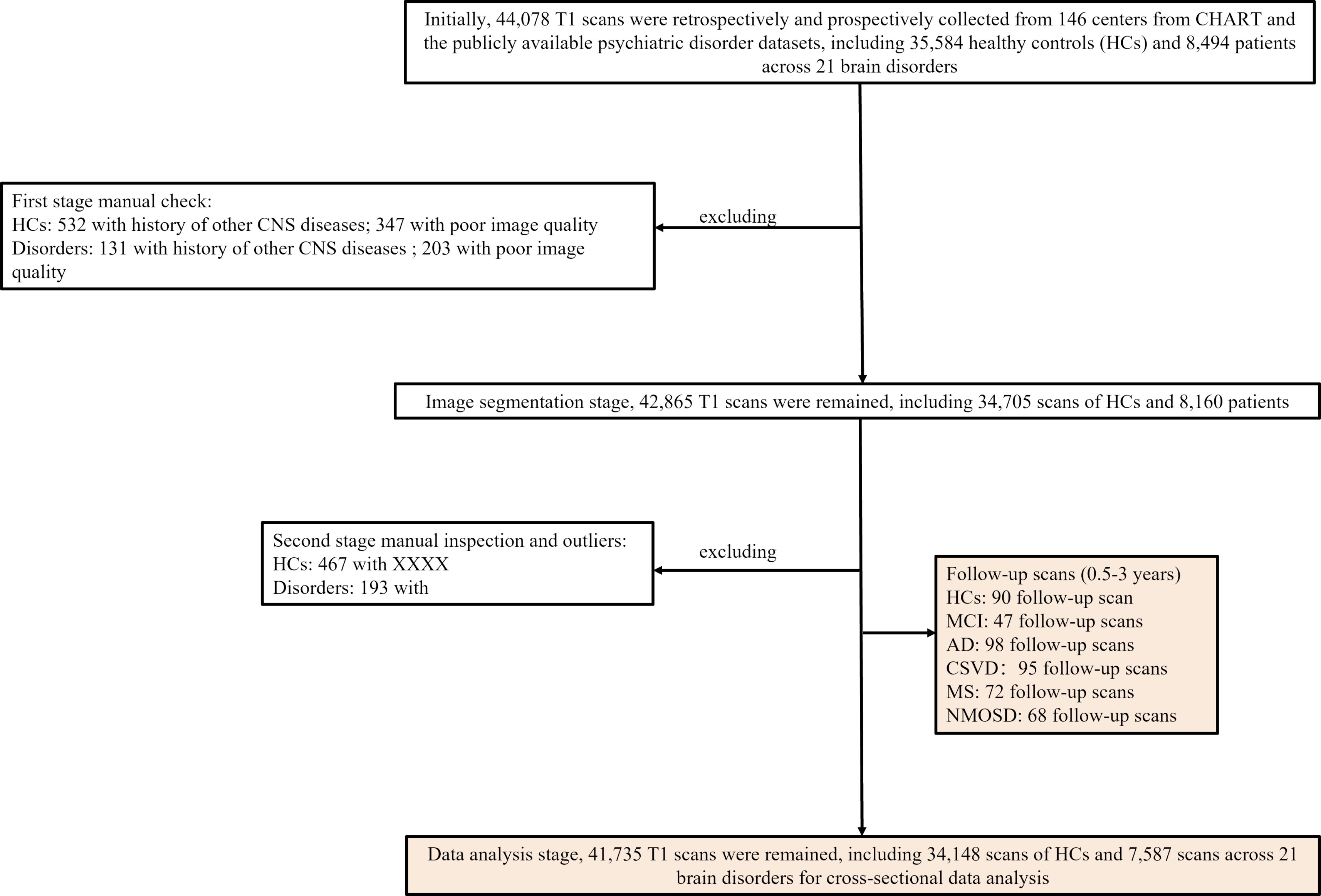
Initially, 44,078 T1 scans were retrospectively and prospectively collected from 146 centers from CHART and the publicly available psychiatric disorder datasets, including 35,584 healthy controls (HCs) and 8,494 patients across 21 brain disorders
excluding
First stage manual check:
HCs: 532 with history of other CNS diseases; 347 with poor image quality
Disorders: 131 with history of other CNS diseases ; 203 with poor image quality
Image segmentation stage, 42,865 T1 scans were remained, including 34,705 scans of HCs and 8,160 patients
excluding
Second stage manual inspection and outliers:
HCs: 467 with XXXX
Disorders: 193 with
Follow-up scans (0.5-3 years)
HCs: 90 follow-up scan
MCI: 47 follow-up scans
AD: 98 follow-up scans
CSVD： 95 follow-up scans
MS: 72 follow-up scans
NMOSD: 68 follow-up scans
Data analysis stage, 41,735 T1 scans were remained, including 34,148 scans of HCs and 7,587 scans across 21 brain disorders for cross-sectional data analysis
