## Extended data Fig.2 for "Unraveling the Lifespan Trajectory of White Matter Hypointensity on T1-weighted MR Images and Clinical Application in Various Brain Disorders"

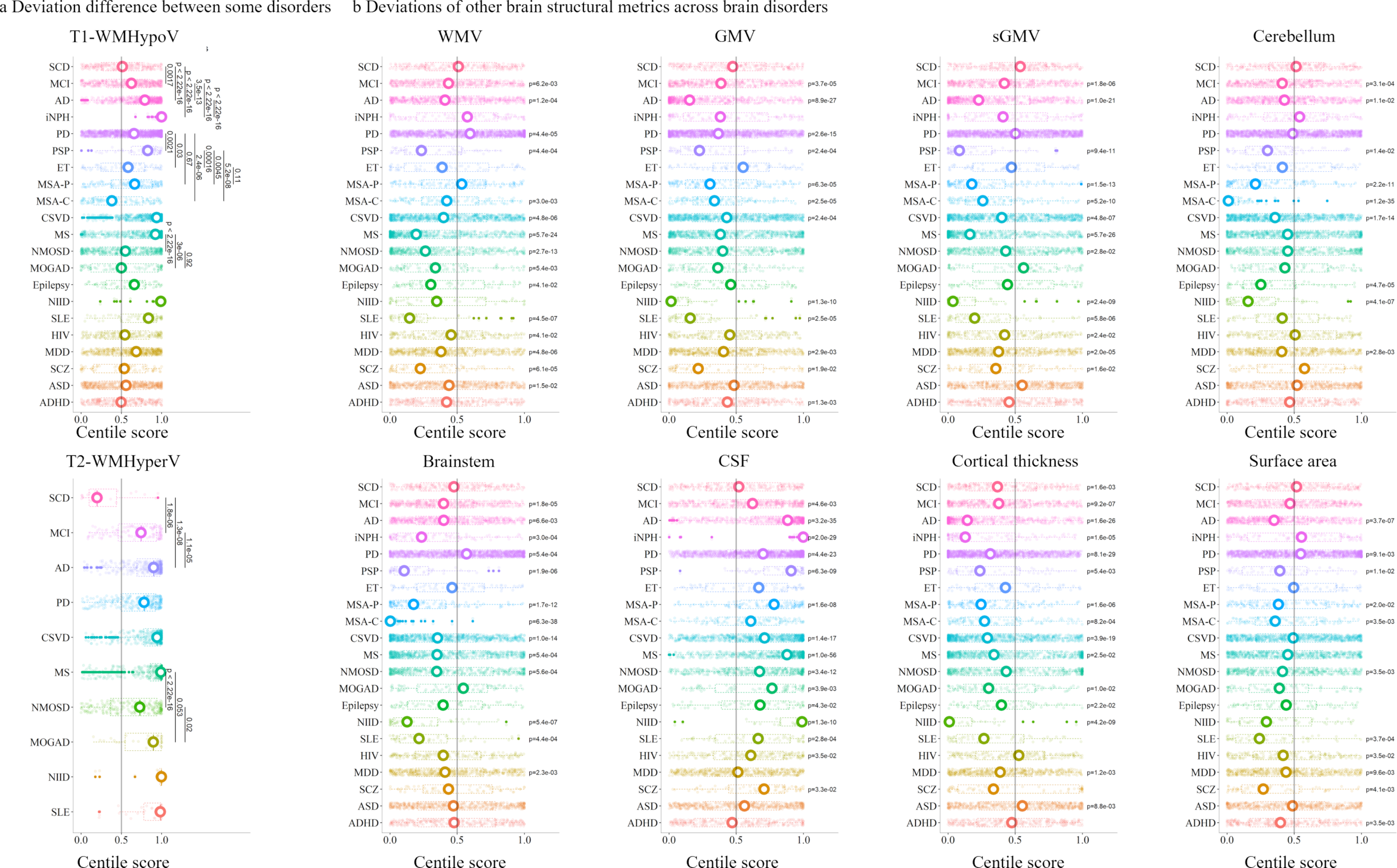

a Deviation difference between some disorders
b Deviations of other brain structural metrics across brain disorders
T1-WMHypoV
SCD
MCI
AD
iNPH
PD
PSP
ET
MSA-P
MSA-C
CSVD
MS
NMOSD
MOGAD
Epilepsy
NIID
SLE
HIV
MDD
SCZ
ASD
ADHD
0.0017
p < 2.22e-16
3.5e-13
p < 2.22e-16
p < 2.22e-16
0.0021
0.03
0.67
2.4e-06
0.00016
0.0045
5.2e-08
0.11
p < 2.22e-16
3e-06
0.92
0.0
0.5
1.0
Centile score
WMV
p=6.2e-03
p=1.2e-04
p=4.4e-05
p=4.4e-04
p=3.0e-03
p=4.8e-06
p=5.7e-24
p=2.7e-13
p=5.4e-03
p=4.1e-02
p=4.5e-07
p=4.1e-02
p=4.8e-06
p=6.1e-05
p=1.5e-02
GMV
p=3.7e-05
p=8.9e-27
p=2.6e-15
p=2.4e-04
p=6.3e-05
p=2.5e-05
p=2.4e-04
p=1.3e-10
p=2.5e-05
p=2.9e-03
p=1.9e-02
p=1.3e-03
sGMV
p=1.8e-06
p=1.0e-21
p=9.4e-11
p=1.5e-13
p=5.2e-10
p=4.8e-07
p=5.7e-26
p=2.8e-02
p=2.4e-09
p=5.8e-06
p=2.4e-02
p=2.0e-05
p=1.6e-02
Cerebellum
p=3.1e-04
p=1.1e-02
p=1.4e-02
p=2.2e-11
p=1.2e-35
p=1.7e-14
p=4.7e-05
p=4.1e-07
p=2.8e-03
T2-WMHyperV
SCD
MCI
AD
PD
CSVD
MS
NMOSD
MOGAD
NIID
SLE
1.8e-06
1.3e-08
1.1e-05
p < 2.22e-16
0.053
0.02
Brainstem
p=1.8e-05
p=6.6e-03
p=3.0e-04
p=5.4e-04
p=1.9e-06
p=1.7e-12
p=6.3e-38
p=1.0e-14
p=5.4e-04
p=5.6e-04
p=5.4e-07
p=4.4e-04
p=2.3e-03
CSF
p=4.6e-03
p=3.2e-35
p=2.0e-29
p=4.4e-23
p=6.3e-09
p=1.6e-08
p=1.4e-17
p=1.0e-56
p=3.4e-12
p=3.9e-03
p=4.3e-02
p=1.3e-10
p=2.8e-04
p=3.5e-02
p=3.3e-02
Cortical thickness
p=1.6e-03
p=9.2e-07
p=1.6e-26
p=1.6e-05
p=8.1e-29
p=5.4e-03
p=1.6e-06
p=8.2e-04
p=3.9e-19
p=2.5e-02
p=1.0e-02
p=2.2e-02
p=4.2e-09
p=1.2e-03
p=8.8e-03
Surface area
p=3.7e-07
p=9.1e-03
p=1.1e-02
p=2.0e-02
p=3.5e-03
p=3.5e-03
p=3.7e-04
p=3.5e-02
p=9.6e-03
p=4.1e-03
p=3.5e-03
Centile score
