## Extended data Fig.3 for "Unraveling the Lifespan Trajectory of White Matter Hypointensity on T1-weighted MR Images and Clinical Application in Various Brain Disorders"

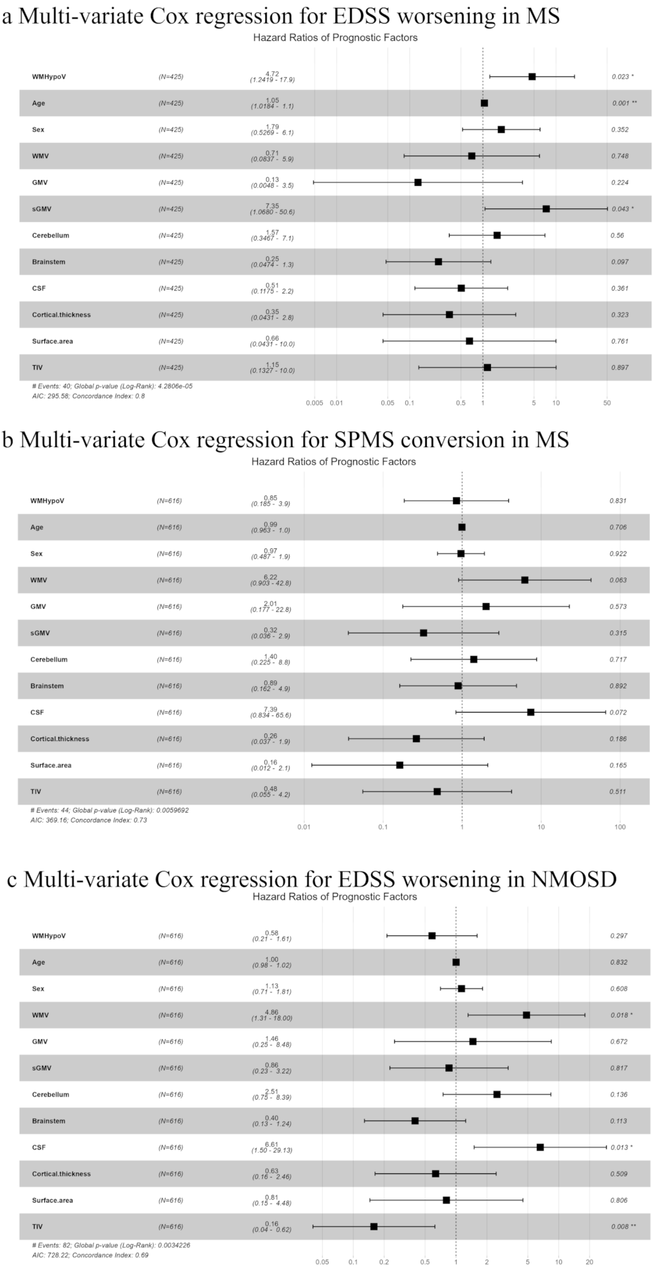
a Multi-variate Cox regression for EDSS worsening in MS
Hazard Ratios of Prognostic Factors
WMHypoV (N=425) 4.72 (1.2419 - 17.9) 0.023 *
Age (N=425) 1.05 (1.0184 - 1.1) 0.001 **
Sex (N=425) 1.79 (0.5269 - 6.1) 0.352
WMV (N=425) 0.71 (0.0837 - 5.9) 0.748
GMV (N=425) 0.13 (0.0048 - 3.5) 0.224
sGMV (N=425) 7.35 (1.0680 - 50.6) 0.043 *
Cerebellum (N=425) 1.57 (0.3467 - 7.1) 0.56
Brainstem (N=425) 0.25 (0.0474 - 1.3) 0.097
CSF (N=425) 0.51 (0.1175 - 2.2) 0.361
Cortical.thickness (N=425) 0.35 (0.0431 - 2.8) 0.323
Surface.area (N=425) 0.66 (0.0431 - 10.0) 0.761
TIV (N=425) 1.15 (0.1327 - 10.0) 0.897
### Events: 40; Global p-value (Log-Rank): 4.2806e-05
AIC: 295.58; Concordance Index: 0.8
0.005 0.01 0.05 0.1 0.5 1 5 10 50
b Multi-variate Cox regression for SPMS conversion in MS
Hazard Ratios of Prognostic Factors
WMHypoV (N=616) 0.85 (0.185 - 3.9) 0.831
Age (N=616) 0.99 (0.963 - 1.0) 0.706
Sex (N=616) 0.97 (0.487 - 1.9) 0.922
WMV (N=616) 6.22 (0.903 - 42.8) 0.063
GMV (N=616) 2.01 (0.177 - 22.8) 0.573
sGMV (N=616) 0.32 (0.036 - 2.9) 0.315
Cerebellum (N=616) 1.40 (0.225 - 8.8) 0.717
Brainstem (N=616) 0.89 (0.162 - 4.9) 0.892
CSF (N=616) 7.39 (0.834 - 65.6) 0.072
Cortical.thickness (N=616) 0.26 (0.037 - 1.9) 0.186
Surface.area (N=616) 0.16 (0.012 - 2.1) 0.165
TIV (N=616) 0.48 (0.055 - 4.2) 0.511
### Events: 44; Global p-value (Log-Rank): 0.0059692
AIC: 369.16; Concordance Index: 0.73
0.01 0.1 1 10 100
c Multi-variate Cox regression for EDSS worsening in NMOSD
Hazard Ratios of Prognostic Factors
WMHypoV (N=616) 0.58 (0.21 - 1.61) 0.297
Age (N=616) 1.00 (0.98 - 1.02) 0.832
Sex (N=616) 1.13 (0.71 - 1.81) 0.608
WMV (N=616) 4.86 (1.31 - 18.00) 0.018 *
GMV (N=616) 1.46 (0.25 - 8.48) 0.672
sGMV (N=616) 0.86 (0.23 - 3.22) 0.817
Cerebellum (N=616) 2.51 (0.75 - 8.39) 0.136
Brainstem (N=616) 0.40 (0.13 - 1.24) 0.113
CSF (N=616) 6.61 (1.50 - 29.13) 0.013 *
Cortical.thickness (N=616) 0.63 (0.16 - 2.46) 0.509
Surface.area (N=616) 0.81 (0.15 - 4.48) 0.806
TIV (N=616) 0.16 (0.04 - 0.62) 0.008 **
### Events: 82; Global p-value (Log-Rank): 0.0034226
AIC: 728.22; Concordance Index: 0.69
0.05 0.1 0.2 0.5 1 2 5 10 20
