## Extended data Fig.4 for "Unraveling the Lifespan Trajectory of White Matter Hypointensity on T1-weighted MR Images and Clinical Application in Various Brain Disorders"

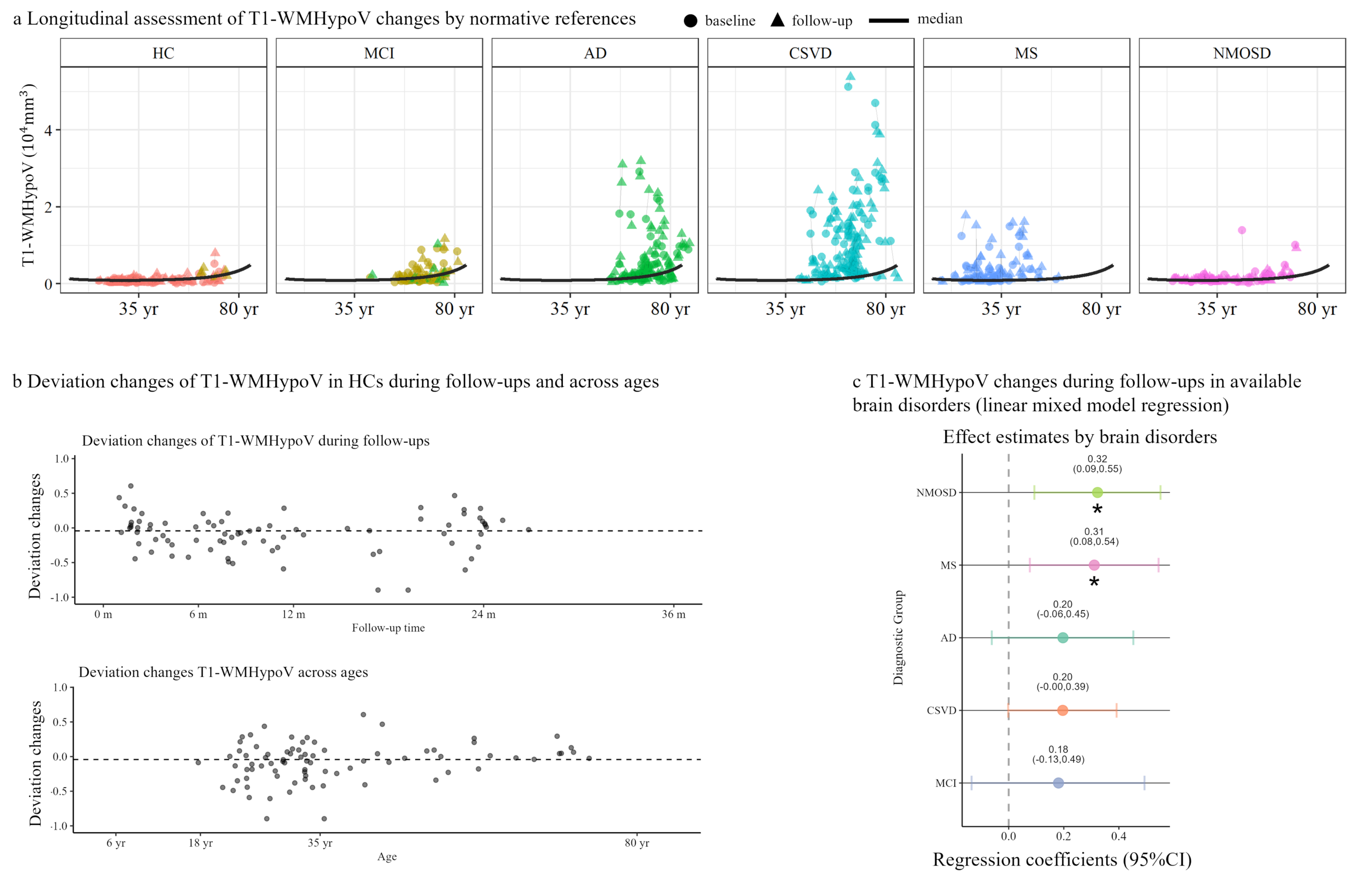
a Longitudinal assessment of T1-WMHypoV changes by normative references
baseline
follow-up
median
HC
MCI
AD
CSVD
MS
NMOSD
T1-WMHypoV (10^4mm^3)
0
2
4
35 yr
80 yr
b Deviation changes of T1-WMHypoV in HCs during follow-ups and across ages
Deviation changes of T1-WMHypoV during follow-ups
Deviation changes
1.0
0.5
0.0
-0.5
-1.0
0 m
6 m
12 m
24 m
36 m
Follow-up time
Deviation changes T1-WMHypoV across ages
6 yr
18 yr
35 yr
80 yr
Age
c T1-WMHypoV changes during follow-ups in available brain disorders (linear mixed model regression)
Effect estimates by brain disorders
Diagnostic Group
NMOSD
0.32
(0.09,0.55)
*
MS
0.31
(0.08,0.54)
*
AD
0.20
(-0.06,0.45)
CSVD
0.20
(-0.00,0.39)
MCI
0.18
(-0.13,0.49)
0.0
0.2
0.4
Regression coefficients (95%CI)
