## Extended data Fig.5 for "Unraveling the Lifespan Trajectory of White Matter Hypointensity on T1-weighted MR Images and Clinical Application in Various Brain Disorders"

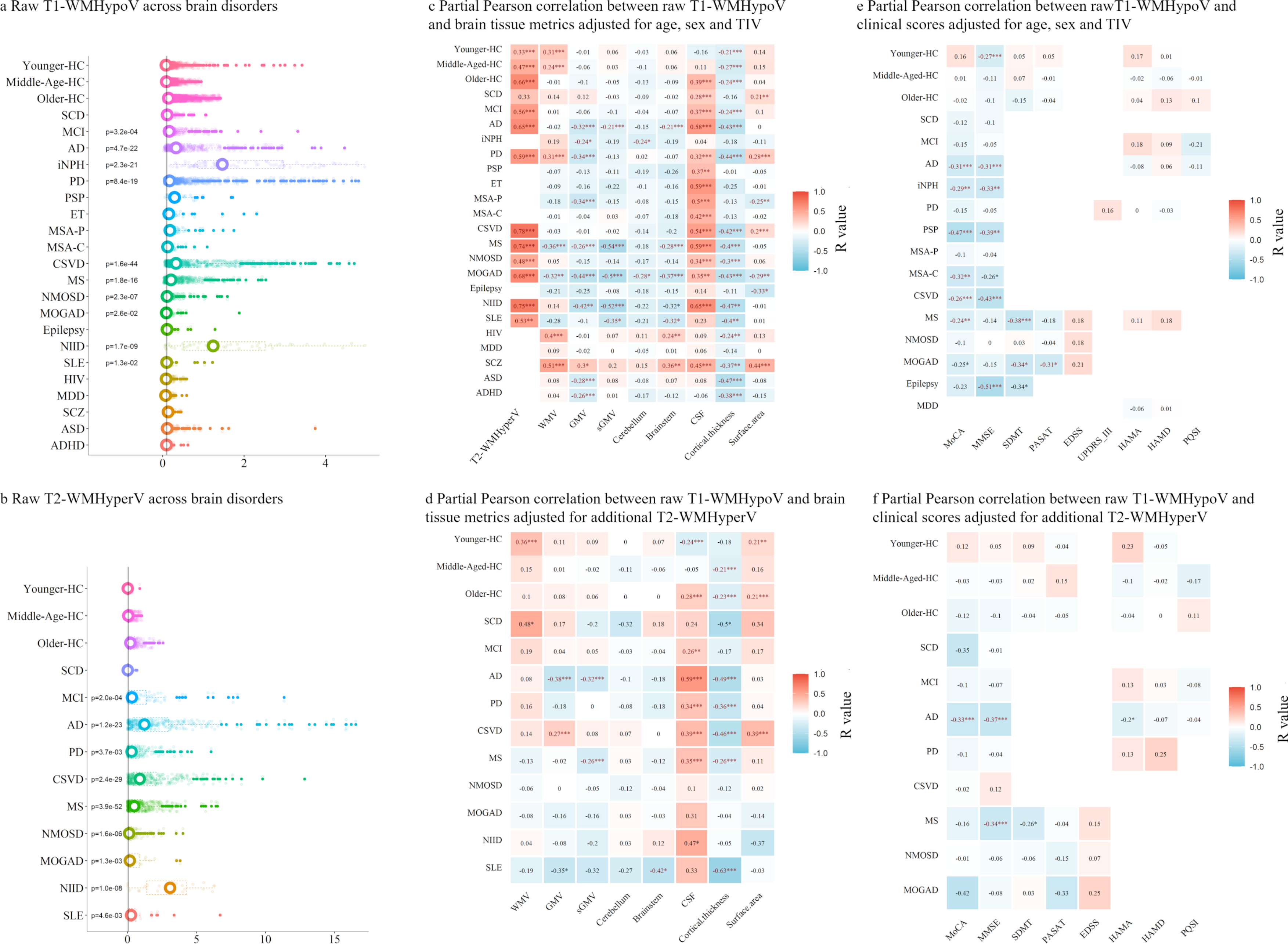
a Raw T1-WMHypoV across brain disorders
Younger-HC
Middle-Age-HC
Older-HC
SCD
MCI
AD
iNPH
PD
PSP
ET
MSA-P
MSA-C
CSVD
MS
NMOSD
MOGAD
Epilepsy
NIID
SLE
HIV
MDD
SCZ
ASD
ADHD
p=3.2e-04
p=4.7e-22
p=2.3e-21
p=8.4e-19
p=1.6e-44
p=1.8e-16
p=2.3e-07
p=2.6e-02
p=1.7e-09
p=1.3e-02
0
2
4
b Raw T2-WMHyperV across brain disorders
Younger-HC
Middle-Age-HC
Older-HC
SCD
MCI
AD
PD
CSVD
MS
NMOSD
MOGAD
NIID
SLE
p=2.0e-04
p=1.2e-23
p=3.7e-03
p=2.4e-29
p=3.9e-52
p=1.6e-06
p=1.3e-03
p=1.0e-08
p=4.6e-03
0
5
10
15
c Partial Pearson correlation between raw T1-WMHypoV and brain tissue metrics adjusted for age, sex and TIV
T2-WMHyperV
WMV
GMV
sGMV
Cerebellum
Brainstem
CSF
Cortical.thickness
Surface.area
R value
1.0
0.5
0.0
-0.5
-1.0
d Partial Pearson correlation between raw T1-WMHypoV and brain tissue metrics adjusted for additional T2-WMHyperV
e Partial Pearson correlation between rawT1-WMHypoV and clinical scores adjusted for age, sex and TIV
MoCA
MMSE
SDMT
PASAT
EDSS
UPDRS_III
HAMA
HAMD
PQSI
f Partial Pearson correlation between raw T1-WMHypoV and clinical scores adjusted for additional T2-WMHyperV
